# Novel *PCDH12* pathogenic missense variants cause neurodevelopmental disorders with ocular malformation

**DOI:** 10.64898/2026.03.05.26343794

**Authors:** Jennifer Rakotomamonjy, Lucas Fares-Taie, Raman Kumar, Cole Gebert, Laura Magaña-Hernandez, Anna Blaszkiewicz, Theresa Benson, Martín Fairbanks-Santana, Angela Trejo, R Curtis Rogers, Claudine Mayer, Kirsley Chennen, Olivier Poch, Tanya M Bardakjian, Thomas F Tropea, Pedro Gonzalez-Alegre, Gemma L Carvill, Jamie Zhang, Shreya Agarwala, Lachlan A Jolly, Nicole J Van Bergen, Shanti Balasubramaniam, Carolyn J Ellaway, John Christodoulou, Jozef Gecz, Jean-Michel Rozet, Alicia Guemez-Gamboa

**Author notes:** Correspondence to: Alicia Guemez-Gamboa, 303 E Chicago Ave, Ward 5-122, CHICAGO IL, 60611, USA. These authors contributed equally to this work (Jennifer Rakotomamonjy, Lucas Fares-Taie, Raman Kumar).

## Abstract

Protocadherin-12 (PCDH12), a cell-adhesion protein belonging to the non-clustered protocadherin family, plays a crucial role in the establishment and regulation of neuronal connections and communication. Bi-allelic loss-of-function (LoF) variants in the *PCDH12* gene have been associated with several neurodevelopmental disorders (NDDs) such as diencephalic-mesencephalic junction dysplasia syndrome, cerebral palsy, and cerebellar ataxia, often accompanied by ocular abnormalities. However, genotypes exhibit variable expressivity. Affected individuals sharing the same *PCDH12* variant presenting differing phenotypic severities have posed major challenges towards identification of the underlying pathogenic mechanisms.

Here, we report three affected individuals from two families, each harbouring non-truncating pathogenic missense variants in *PCDH12*. The patients are compound heterozygous, with each individual carrying one extracellular [c.1742T>G (p.Val581Gly) and c.1861_2del/insCA (p.Ile621His)] and one intracellular variant [c.3370C>T (p.Arg1124Cys) and c.3445G>A (p.Asp1149Asn] on each allele. The children present with a range of phenotypes similar to those associated with LoF variants. One child exhibited microcephaly and seizures, while the two siblings displayed developmental delays and severe behavioral disorders. All three children experienced some degree of visual impairment.

The missense variants provided new insights into the neurodevelopmental consequences of compromised *PCDH12* function by distinguishing the specific consequences associated with dysfunction in the extracellular versus intracellular domains of PCDH12. All identified missense variants are predicted to be deleterious and destabilizing. The expression of PCDH12 in HEK293T and HeLa cells demonstrated that PCDH12 is expressed effectively, regardless of the presence of missense variants. However, the extracellular variants p.Val581Gly and p.Ile621His compromised the stability of PCDH12’s homophilic adhesion. Additionally, we found evidence of an interaction between PCDH12 and the extracellular domain of the epilepsy-associated PCDH19 protein. PCDH12 extracellular missense variants also affect PCDH19 stability.

Our study provides evidence that PCDH12 mediates both homophilic and heterophilic interactions. Our findings also highlight the importance of stable PCDH12-mediated adhesion, emphasizing the need to further study the functional consequences of *PCDH12* missense variants on brain and visual system development.

## Introduction

Protocadherins (PCDHs) are a diverse subfamily of cell adhesion molecules within the cadherin superfamily, playing pivotal roles in cell recognition, neuronal migration, and differentiation.^1^ Among them, *PCDH12*, a member of the non-clustered PCDH family, encodes a membrane-bound glycoprotein predominantly expressed in the developing brain and vasculature.^2,3^ Genetic studies have identified biallelic LoF pathogenic variants in *PCDH12* as contributing factors to various neurodevelopmental disorders (NDDs). These disorders include developmental delay, movement disorders, epilepsy, and structural brain abnormalities such as microcephaly, midbrain malformations, and intracranial calcification.^3–12^ Interestingly, *PCDH12*-associated disorders are known to display variability in clinical expressivity, with affected individuals carrying the same *PCDH12* variants exhibiting varying phenotypic severity (**Supplementary Table 1**).^7,9,11,12^

Given that disruptions in brain development often manifest as significant structural and functional anomalies, it is crucial to investigate the role of PCDH12 as a cell adhesion molecule that is important for neuronal wiring to better understand the disease pathology. The existing evidence suggests that PCDH12 is integral to maintaining neural architecture and ensuring proper connectivity within neural circuits. Experimental studies using cortical organoid models have demonstrated that loss of *PCDH12* can prematurely drive neuronal differentiation and disrupt migration, further highlighting its significance in brain development.^13^

In addition to its established role in the central nervous system, PCDH12 disruption is associated with a range of ocular anomalies, including cataracts, retinal dystrophies, and various structural eye defects.^4,7–10,12,14^ The shared developmental pathways between the brain and eye indicate that variants in PCDHs, such as PCDH12, can affect both systems simultaneously. This observation is important because patients with *PCDH12* variants exhibit both neurodevelopmental and ocular phenotypes, underscoring the gene’s essential role in coordinating the developmental processes of both the brain and visual system.

Here, we report the identification of the first non-truncating compound heterozygous missense variants in *PCDH12* in patients diagnosed with NDDs. Interestingly, these patients exhibit intellectual disability and delayed psychomotor development with one of them exhibiting microcephaly. This observation suggests that these missense variants may lead to loss of PCDH12 function similar to the effect of the protein truncating mutations, offering valuable insights into PCDH12 function at the protein domain level.

We have used a combination of genomics, *in silico* modeling, and *in vitro* assays to uncover whether these newly identified missense variants affect PCDH12 adhesive function and interaction with the actin cytoskeleton.

## Materials and methods

### Human studies

Affected individuals were identified from genetics and/or neurology clinics targeting patients with NDDs. Identification of eligible patients was facilitated through personal communication and the GeneMatcher platform^15^. All enrolled patients and their families participated under Institutional Review Board- or Human Research Ethics Committees-approved protocols and provided informed consent for their inclusion in the study. Genomic sequencing was performed on one or two affected individuals or on a familial trio consisting of the parents and one affected child from each family. Exome sequencing was carried out by a certified commercial clinical laboratory located in the United States (Family 3) and by the Genomic Core Facility of Imagine (Paris, France) (Family 4). Details pertaining to each patient, including their clinical phenotype, are summarized in **Table 1**.

**Table 1.**
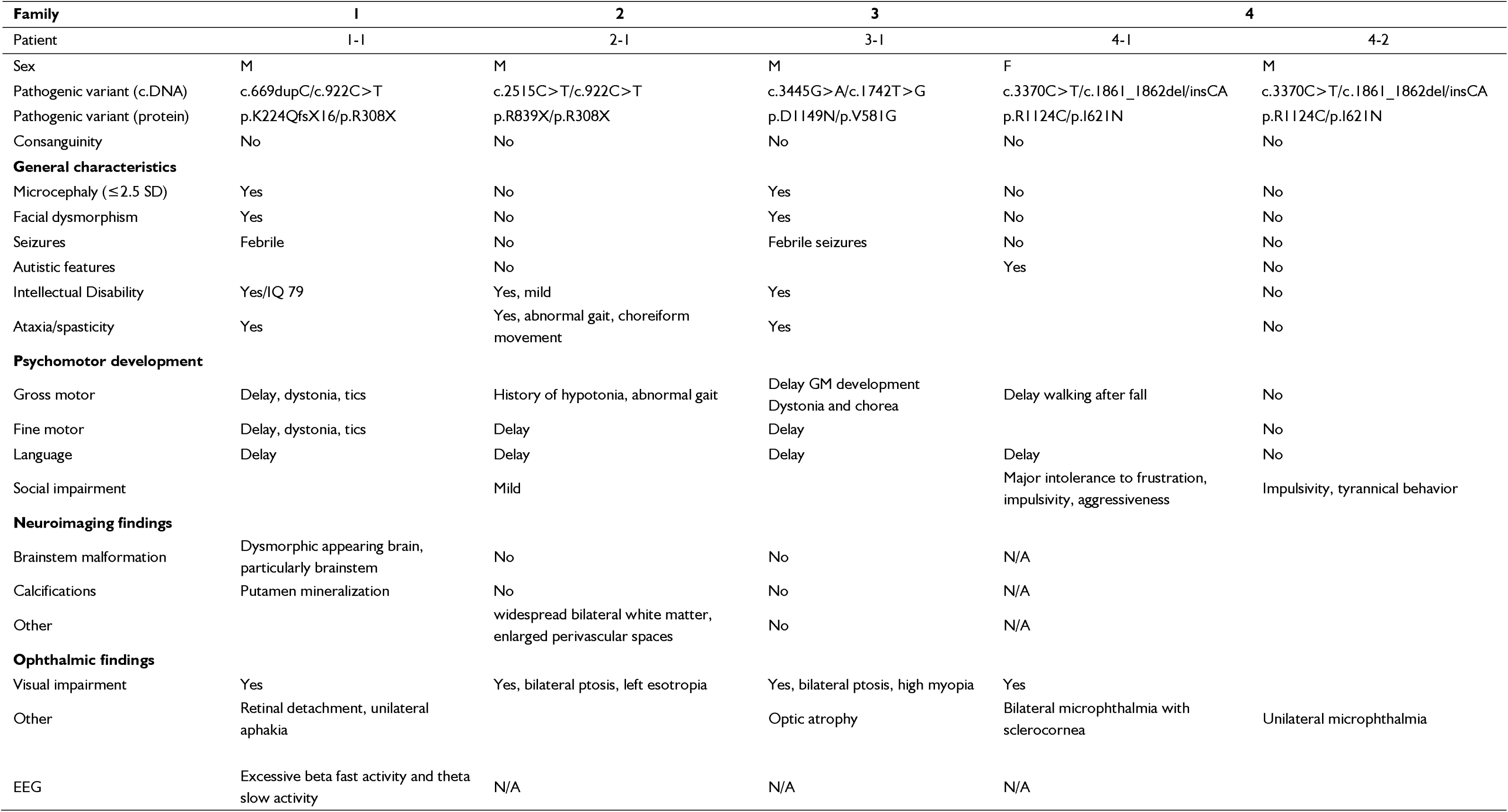
Main clinical features of affected individuals with biallelic heterozygous PCDH12 variants.

### Exome sequencing analysis

Trio (Family 3) and whole (Family 4) exome sequencing and bioinformatic analyses for the probands and their parents were performed as previously described.^16,17^ Variants were filtered under both recessive and dominant inheritance models, including consideration of *de novo* events and incomplete penetrance, to identify disease-causing variants.

### Variant analysis

The variants were annotated with the Ensembl VEP program^18^ in association with the dbNSFP database (v4.4)^19,20^ to extract different deleteriousness predictor scores (**Supplementary Table 2**). The pathogenicity of the selected variants was evaluated using the Alamut Mutation Interpretation Software (http://www.interactive-biosoftware.com), a comprehensive decision support system that integrates multiple tools for mutation analysis, including Align GVGD, MutationTaster, PolyPhen-2, SIFT, SpliceSiteFinder-like, MaxEntScan, NNSPLICE, GeneSplicer, Human Splicing Finder, ESEfinder, and RESCUE-ESE. Co-segregation of the identified *PCDH12* variants with the disease was confirmed by Sanger sequencing in all available family members.

### Sequence analysis

Sequences belonging to the δ2-protocadherin family (PCDH8, PCDH10, PCDH12, PCDH17, PCDH18 and PCDH19)^21^ of 20 phylogenetically representative vertebrates were retrieved from Uniprot Database^22^ or NCBI Database^23^, when absent in Uniprot. The 134 retrieved sequences were aligned with Pipealign2^24^ and manually corrected. A phylogenetic tree of the full-length and on the 6-cadherins extracellular (EC) domain sequences with bootstrap NJ method (number of bootstrap trials of 1000) were drawn using ClustalX.^25^ The mean percentage identity of each individual cadherin module was calculated for the 6 PCDH proteins.

### Structural analysis

The different 3D structure models of human PCDH8 (O95206), PCDH10 (Q9P2E7), PCDH12 (Q9NPG4), PCDH17 (O14917), PCDH18 (Q9HCL0), and PCDH19 (Q8TAB3) were retrieved using the neural network-based Alphafold2 tool.^26^ and were predicted by using a large language model approach via a local instance of ESMFold^27^ with default parameters.

As PCDHs contain an extra- and an intracellular domain with an extended domain organization, we manually corrected the model by moving away the extracellular part to propose a more reasonable conformational state. The different models were then superimposed and compared. The 6 cadherin-module boundaries could be defined in ESM vs 18 % in AF2 model for PCDH12. DDMut was used to determine the effect of the mutation on the structure.^28^

### In vitro studies

#### Plasmid constructs and generation of *PCDH12* missense variants by Site-directed mutagenesis

The PCDH15-Fc (PCDH15 extracellular domain fused to the Fc domain of immunoglobulins) and pCDNA3-Fc plasmids^29^ were kindly provided by the Müller Lab. The extracellular domain of human PCDH12 (hPCDH12), featuring *Hin*dIII overhangs, was amplified via PCR. The PCDH15 sequence in PCDH15-Fc was replaced with this fragment to generate the hPCDH12-Fc plasmid. The following primers were used for amplification: 5’- GGTGGTAAGCTTATGATGCAACTTCTGCAA-3’; 5’-GGTGGTAAGCTTGAGCTGAGTCCCTCAGGTGGT-3’ of the PCDH12 fragment. The full-length hPCDH12 plasmid containing eGFP or mCherry were generated using a Gateway-compatible destination vector pmax-mCherry generously provided by the Maniatis lab.^30^ The full-length hPCDH12 sequence was initially cloned into the donor plasmid pDONR221, followed by Gateway cloning to produce the PCDH12-mCherry expression vector. For the eGFP constructs, the mCherry reporter was replaced with eGFP, and Gateway cloning was performed to integrate hPCDH12. The following Gateway cloning primers were used for amplifying the full-length hPCDH12 sequence: 5’-GGGGACAAGTTTGTACAAAAAAGCAGGCTGCATGATGCAACTTCTGCAA-3’; 5’-GGGGACCACTTTGTACAAGAAAGTTGGGCACAGGCACCTGCTGCTGCT-3’. Patient-derived *PCDH12* variants c.1742T>G (p.Val581Gly), c.1861_1862del/insCA (p.Ile621His), c.3370C>T (p.Arg1124Cys), and c.3445G>A (p.Asp1149Asn) were introduced into the human full-length hPCDH12 plasmids (eGFP and mCherry) via site-directed mutagenesis using high-fidelity PCR amplification (Phusion DNA polymerase). Additionally, the c.1742T>G (p.Val581Gly) and c.1861_1862del/insCA (p.Ile621His) variants were incorporated into the hPCDH12-Fc plasmid using the same method.

Following amplification, template DNA was eliminated by digestion with the methylase-sensitive *Dpn*I restriction enzyme for 1 hour at 37°C. XL1-Blue competent bacteria (Agilent Technologies) were transformed using the digested products and plated on LB Agar supplemented with ampicillin (100 μg/mL). The variants from single colony DNA minipreps were validated using whole plasmid sequencing (Plasmidsaurus).

#### Cell cultures

HEK293T and HeLa cells were cultured in Dulbecco’s modified Eagle medium (DMEM) with high glucose, L-Glutamine and sodium pyruvate (Gibco), supplemented with 10% fetal bovine serum (Gibco). Cells were grown at 37°C in 5% CO_2_.

#### Expression constructs and transfections

For the immunofluorescence experiments, 75,000 HEK293T cells per well of a 12-well plate were plated onto glass coverslips coated with poly-L-ornithine and laminin one day prior to transfection. Cells were then transfected with 1 µg of constructs expressing mCherry or GFP-fused WT or variant *PCDH12* using Lipofectamine 2000 (Invitrogen-ThermoFisher Scientific).

For the western blot experiments, HEK293T cells were plated at a density of 1.5×10^5^ cells/well of a 6-well plate, 48 hours prior to transfection. Cells were then transfected with 2.5 µg of constructs described above using Lipofectamine 2000 (Invitrogen-ThermoFisher Scientific).

For the bead aggregation assays, HEK293T cells were plated at a density of 3×10^5^cells per 10-cm dish 48 hours prior to transfection. Cells were then transfected with 17.3 µg of Fc-fused WT or variant PCDH12-ectodomain constructs using Lipofectamine 3000 (Invitrogen-ThermoFisher Scientific).

For the co-immunoprecipitation assays, HEK293T cells were plated at 5×10^5^ cells/well in 6-well culture plates. The next day, cells were transfected with one or more expression plasmids using Lipofectamine 3000 reagent and harvested 24 hours post-transfection. First, the cells were lysed in cold lysis buffer (50 mM Tris-HCl pH 7.5, 150 mM NaCl, 0.4% Triton-X-100, 2 mM EDTA, 0.01% SDS, 50 mM NaF, 0.1 mM Na_3_VO_4_ and 1× Protease inhibitor/no EDTA, Merck) and sonicated at 25% Amplitude for 15 seconds on Sonic’s Vibra-Cell VCX. The lysates were centrifuged at 15,000 rpm for 30 minutes at 4°C, following which 10% clarified lysate was saved as input, and the remaining lysate was subjected to immunoprecipitation using anti-c-Myc-Magnetic Dynabeads (Thermo Fisher) as previously reported^31^. Immunoprecipitated complexes were eluted using 1× SDS protein loading buffer (62.5 mM Tris-HCl, pH 6.8, 2% SDS, 10% glycerol, 5% β-mercaptoethanol) and stored at -80°C until western blot analyses. Each experiment was replicated twice.

#### Immunofluorescence and imaging

24-hour post-transfection, HEK293T cells were fixed in ice-cold 4% paraformaldehyde for 15 minutes, followed by three washes with 1X PBS. Cells were then incubated in a PBS-based blocking buffer containing 0.3% Triton X-100 and 10% normal donkey serum (NDS). HEK293T cells were labeled with a mouse primary antibody anti-WAVE1 (Millipore) in 1X PBS containing 0.1% Triton X-100 and 5% NDS overnight at 4°C. After three additional washes with 1X PBS, cells were incubated with a donkey secondary antibody anti-mouse Alexa647 (Invitrogen-ThermoFisher Scientific), diluted in 1X PBS for two hours at room temperature. Following further washes, HEK293T cells were stained with Hoechst 33342 for 10 minutes at room temperature. Finally, the coverslips were mounted onto microscopy glass slides using Fluoromount G (Southern Biotech). Images were acquired using a Nikon A1 confocal laser microscope system with x63 or x100 magnification oil immersion lenses at 1024×1024 pixel resolution.

#### Protein expression

24-hour post-transfection, HEK293T cells were lysed in 1X RIPA buffer (Millipore) supplemented with a protease and phosphatase inhibitor cocktail (ThermoFisher Scientific). Protein concentrations were determined by BCA assay (Pierce), and 30 μg of total protein was resolved by SDS-PAGE before being transferred onto PVDF membranes. The membranes were blocked with Intercept (TBS) Blocking Buffer (LICORbio) and then incubated with primary antibodies - rabbit anti-mCherry (Abcam), chicken anti-GFP (Abcam), and mouse anti-α-Tubulin (Abcam) - diluted in Intercept containing 0.2% Tween20 overnight at 4°C with gentle agitation. The following day, the membranes were incubated for two hours at room temperature with secondary antibodies - IRDye 680RD Goat anti-Rabbit IgG, IRDye 680RD Goat anti-Mouse IgG, and IRDye 800CW Donkey anti-Chicken - diluted in Intercept supplemented with 0.2% Tween20 and 0.01% SDS. The blots were imaged using the Odyssey Fc Imager (Li-Cor), and quantification was performed using Image Studio™ software (Li-Cor).

For the co-immunoprecipitation experiments, input and immunoprecipitated samples were analyzed by western blotting. Proteins were resolved in 7% homemade SDS-PAGE gels and transferred onto nitrocellulose membrane. Following blocking the membranes in 10% skim milk in 1× Tris-buffered saline, 0.1% Tween 20 (1× TBST), they were probed with mouse-anti-Myc-HRP (1:5000) or primary mouse-anti-HA (400ng/ml) overnight at 4°C and secondary antibody (for HA-tagged proteins), goat-anti-mouse-HRP (1:1000) for 2 hours at room temperature in 2% skim milk in 1× TBST. The membranes were washed with 1× TBST and target proteins visualized using Clarity Western ECL substrate (Bio-Rad) on ChemiDoc XRS+ System (Bio-Rad).

#### Bead aggregation assay

The day following transfection, HEK293T cells were rinsed twice with DMEM and then returned to the incubator for an hour. After which, the cells were rinsed a third time with DMEM. HEK293T cells were left to produce Fc-fusion proteins for 48 hours before the media was collected. Upon collection, the cell media were centrifuged at 500x *g* for 5 minutes to pellet the cell debris, followed by filtration through a 0.45-μm syringe filter into a 10 kDa molecular weight cut-off Ultra centrifugal filter (Amicon, Millipore-Sigma). The Ultra centrifugal filters were then spun down at 4,000x *g* for 15 minutes at 4°C to yield approximately 500 μL of concentrated media. The Fc-fused proteins were bound to 1 µL of magnetic green (screenMAG/G; Chemicell) or red protein-A beads (screenMAG/O; Chemicell) for 2 hours at 4°C with gentle rotation in 1 mL of ice-cold Binding Buffer (50 mM Tris, 100 mM NaCl, 10 mM KCl, and 0.2% BSA, pH 7.4). Beads were then quickly washed twice with 1 mL of ice-cold Binding Buffer and then resuspended in 300 µL of the same buffer. Resuspended beads were split into two equal portions of 150 µL, placed into low-attachment 24-well plates, and treated with 1.5 µL of 200 mM CaCl_2_ to induce adhesion. Images were collected using a BZ-X710 inverted fluorescence microscope (Keyence) after 30 minutes incubation at room temperature. Aggregate sizes were assessed using ImageJ. A cutoff of 5 µm^2^ was used to discriminate between aggregated and non-aggregated beads, as it was the average size of control Fc-bound beads (4.96 ± 0.88 µm^2^). To quantify size distribution, aaggregates were ranked into incremental 5-µm^2^ bins.

#### Transactivation and RNAseq

As *PCDH12* gene is very lowly expressed in human fibroblasts, we used our gene transactivation protocol to induce PCDH12 expression^32^ and performed RNA-sequencing to determine if the *PCDH12* transcripts in the fibroblasts carrying the c.669dup (p.Lys224Glnfs*16) or c.922C>T (p.Arg308*) variants were spliced correctly and/or degraded via nonsense-mediated mRNA decay. The *PCDH12* gene transactivation kit was generated as previously described using the following *PCDH12* specific guide RNAs (PCDH12_gRNA_1: GCCCTCCCTTTTCCTGTAGG; PCDH12_gRNA_2: GCCTCATCCTCCTCAGCGGC; PCDH12_gRNA_3: GTCCACGTGCAAGCAGGTGG and PCDH12_gRNA_4: GACAGAGAGGTAAACAAGAT). The control (n=4) and variant fibroblasts were transactivated and cultured in the presence or absence of 200 μg/mL cycloheximide (CHX) (Sigma-Aldrich) for 24 h before harvesting the cells for RNA extraction and RNAseq as reported^32^. Strand specific RNA-seq was performed using the MGI T7 PE150 sequencer at 100M total reads per sample (Biomarker Technologies Company Limited, Hong Kong) and data analysed as previously described.^32^ Transactivation resulted in robust *PCDH12* expression in control and variant fibroblasts, averaging 245 ± 75.5 transcripts per million (± standard error of mean) across n=14 experiments.

### Statistical analysis

Statistical analyses were performed using GraphPad prism 10. After confirming normality using the Shapiro-Wilk test, mCherry relative expression in HEK293T cells was analyzed with an ordinary one-way ANOVA followed by Holm-Šídák multiple comparisons test. Bead aggregation data were analyzed using the Brown-Forsythe and Welch ANOVA tests followed by the Dunnett’s T3 multiple comparisons test (**Fig. 4B**). In case of violation of normality, the Kruskall-Wallis test was used followed by the Dunn’s multiple comparisons test (**Supplementary Fig. 3B**). For the comparison of bin distribution in the bead aggregation experiments, the Fisher’s exact test was used (**Fig. 4D-F**; **Supplementary Fig. 3C-E**). The level of significance was set at *P* < 0.05 for all statistical tests.

## Results

### Novel *PCDH12* pathogenic missense variants result in NDDs with ocular malformations

To date, all identified homozygous or compound heterozygous *PCDH12* variants linked to NDDs are predicted to result in protein loss (**Supplementary Table 1**).^3,4,6–12^ Analysis of the 39 previously published cases reveals that *PCDH12*-related variants display variable clinical expressivity; individuals with the same *PCDH12* variant, even within the same family, exhibit differing levels of severity and/or types of clinical features (**Supplementary Table 1**). This variability poses challenges in identifying the specific mechanisms underlying the disease and highlights the necessity of expanding the patient population to enhance our understanding of *PCDH12*-related pathology. Using GeneMatcher,^15^ we found five additional individuals from four unrelated families with bi-allelic *PCDH12* variants. Two of the affected individuals harbored truncating variants in a compound heterozygous state, including the previously reported c.669dup (p.Lys224Glnfs*16)^8^ (Family 1) and c.2515C>T (p.Arg839*)^4^ (Family 2) variants, as well as a novel nonsense substitution, c.922C>T (p.Arg308*) (Family 1 and 2) (**Fig. 1A**, **Table 1**). This latter is an ultra-rare variant in gnomAD v4.1.0 (rs370283860, allele frequency: *AF* = 0.00006196) **(Supplementary Table 2**). Consistent with previous findings, the affected individuals exhibited a complex NDD in the setting of microcephaly, intellectual disability, general developmental delay, and varying degrees of visual impairment (**Table 1**). To assess the effects of truncating variants on *PCDH12* mRNA processing, we performed gene transactivation^32^ to induce the expression of *PCDH12* in human dermal fibroblasts collected from healthy individuals (n=4) or individuals with truncating variants (n=3), including the individual from Family 2. Transactivated cells were cultured in the presence or absence of CHX to assess regulation by nonsense mediated mRNA decay and isolated mRNA subjected to RNA sequencing.^32^ Transactivation culminated in robust *PCDH12* mRNA expression across all samples (average transcripts per million (TPM) ± standard error of the mean = 244.7 ± 75). There was no major alteration in the splicing of *PCDH12* variant mRNAs compared to controls (**Supplementary Fig**. **1A**). Treatment of samples with CHX unveiled the inclusion of a novel poison exon derived from intron 2 in a proportion of mRNAs isolated from both controls and variant samples (**Supplementary Fig**. **1A**). While on average, the abundance of *PCDH12* mRNA increased 2-fold in control samples following CHX treatment, this increase was significantly higher (∼4-fold, *p*<0.05, Student’s t-test) in individuals with truncating variants aligned with heightened decay of truncating variant mRNAs by NMD and LoF mechanism of action (**Supplementary Fig. 1B**). Although these variant mRNAs undergo NMD, our assay shows that mRNA decay is not complete, allowing a significant number of variant mRNAs to escape NMD. While this may be an artifact of the overexpression in fibroblasts, if it is physiologically relevant, we can speculate that varying levels of escape from NMD in different individuals or tissues could contribute to differences in phenotypes, especially if the escaping mRNAs are translated into truncated peptides.

**Figure 1.**
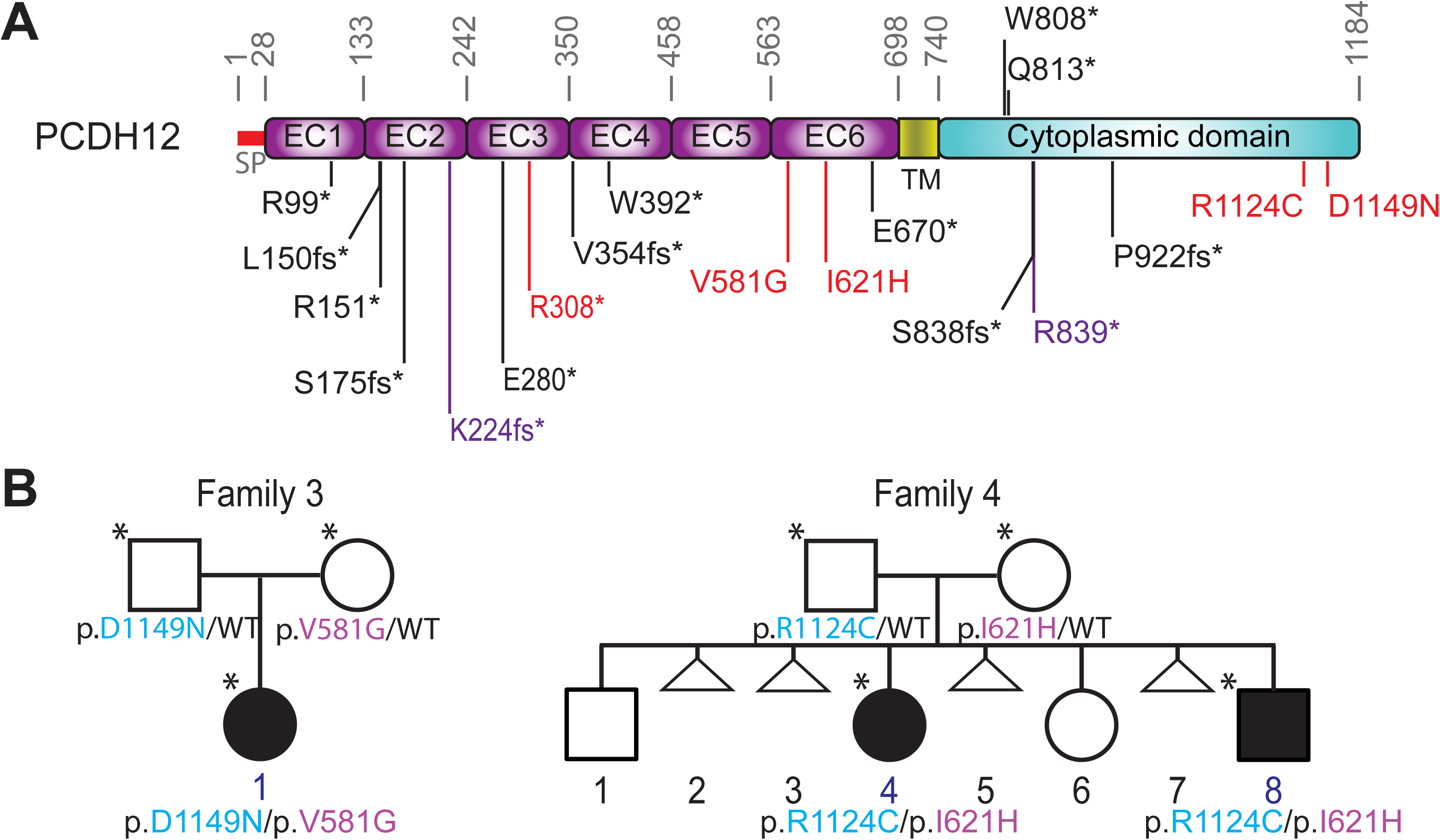
Bi-allelic pathogenic variants in *PCDH12* lead to neurodevelopmental phenotypes. (**A**) Location of pathogenic variants relative to predicted protein. Pathogenic variants previously identified (black), newly reported in this study (red), or both (purple) are shown. (**B**) Pedigrees of non-consanguineous families with compound heterozygous missense *PCDH12* pathogenic variants. SP=Signal Peptide; EC=Extracellular Cadherin domain; TM=Transmembrane domain.

In addition to these truncating variants, we describe compound heterozygous *PCDH12* missense variants in the remaining two families including c.1742T>G (p.Val581Gly) and c.3445G>A (p.Asp1149Asn), and c.1861_1862del/insCA (p.Ile621His) and c.3370C>T (p.Arg1124Cys) (**Fig. 1A**). In the first family, the *PCDH12* variants c.1742T>G (p.Val581Gly) and c.3445G>A (p.Asp1149Asn) were identified by trio-exome sequencing in a preteen male born to non-consanguineous parents (Family 3; **Fig. 1A, B**). He presented with developmental delay from infancy and deteriorated clinically, with profound intellectual disability, progressive microcephaly, bilateral ptosis, high myopia, and optic atrophy causing severe visual impairment. In addition, he exhibited chorea and dystonia. Brain MRI scan revealed no brain structural anomalies (**Table 1**). Neither the c.1742T>G nor the c.3445G>A variant is present in existing population-scale genetic databases. The c.1742T>G, p.Val581Gly substitution changes the highly conserved valine 581 in the EC6 domain to a glycine (p.Val581Gly), which is predicted to be highly deleterious (*Grantham distance*: 109 [0-215]). The c.3445G>A, p.Asp1149Asn variant alters a highly conserved aspartic acid at position 1149 to an uncharged polar asparagine with most *in silico* pathogenicity predictors assessing this variant as deleterious (*Grantham distance*: 23 [0-215]) (**Supplementary Table 2**).

The *PCDH12* variants c.1861_1862del/insCA (p.Ile621His) and c.3370C>T (p.Arg1124Cys) were identified by analysis of exome sequencing datasets from two affected siblings (sister and brother) and their unrelated parents (Family 4; **Fig. 1B**). The affected sister was born blind due to severe microphthalmia and sclerocornea affecting both of her eyes. She exhibited delayed walking and verbal communication. She was diagnosed with severe autism following concerns about her social and learning abilities. She also exhibited severe behavioral problems, including impulsivity, agitation, and aggression. Her behavioral issues complicated medical imaging studies, including cerebral MRI.

The affected brother suffers from unilateral severe microphthalmia in his right eye. He displays impulsive and tyrannical behaviors. These behavioral issues manifest as major intolerance to frustration, aggressiveness, and incidents of hitting and biting. Behavioral difficulties hampered cerebral MRI. His neuropsychomotor development appears to be normal for his age. The c.1861_1862delinsCA (p.Ile621His) variant is not detected in population-scale genetic databases. This complex indel variant leads to a single amino acid substitution (p.Ile621His) which affects a highly conserved, uncharged hydrophobic isoleucine in the cadherin domain into a positively charged histidine (p.Ile621His; *Grantham distance*: 94 [0-215]). The c.3370C>T substitution is a known ultra-rare variant (rs778087717, *AF* = 0.00002354) which replaces a positively charged Arginine in the C-terminal portion of PCDH12 with a sulfur-containing Cysteine (p.Arg1124Cys; *Grantham distance*: 180 [0-215]). Both variants are predicted to be deleterious by most *in silico* pathogenicity predictors (**Supplementary Table 2**). Sanger sequencing analysis of all members of the family confirmed the co-segregation of the disease with the *PCDH12* variants by showing single heterozygosity for either variant in the mother’s and father’s DNA, respectively, and absence or presence of one allele only in the unaffected brother and sister, respectively (**Fig. 1B**). Notably, these four residues are located in highly conserved subregions across species, with GERP score > 3 (**Supplementary Fig. 1C, Supplementary Table 2**).

### Novel *PCDH12* extracellular pathogenic missense variants likely result in loss of function

These new missense variants were identified in individuals with complex NDDs, which resemble the patients carrying homozygous truncating variants. This presented a unique opportunity to explore the functions of PCDH12 subdomains. PCHD12 is a member of the non-clustered δ2-PCDH subfamily (that includes PCDH8, PCDH10, PCDH17, PCDH18, and PCDH19). All PCDHs have an extracellular ectodomain consisting of six (α, β, γ, and δ2-PCDHs) or seven (δ1-PCDHs) cadherin motifs and a variable cytoplasmic domain. δ-PCDH intracellular domain harbors two (δ2-PCDHs) or three (δ1-PCDHs) <u>C</u>onserved <u>M</u>otifs (CM1-3). PCDH12 is unique as it does not share these CMs, harboring only a <u>W</u>ave <u>R</u>egulatory <u>C</u>omplex (WRC) binding motif that mediates interaction with the WRC and the actin cytoskeleton. This conserved sequence motif is also present in clustered α-PCDHs and in all other δ2-PCDHs.^33^ The sequence and phylogenetic analyses confirm that PCDH17, PCDH19, and PCDH10 are closely related while PCDH12 is close to PCDH18 (and PCDH8 being more distant) when considering the full-length sequences (**Supplementary Fig. 1D)** as well as the extracellular domain alone (not shown). Mean percentage identity of the cadherin repeat reveals that the fourth module (EC4) is the most conserved in PCDH8, PCDH17, PCDH18, and PCDH19 while EC2 is the most conserved in PCDH10 and 12. Interestingly, although EC6 is the least conserved cadherin repeat in PCDH12, as well as in PCDH8 and PCDH18 (**Fig. 2A**), the patient-specific PCDH12 variants at Val581 and Ile621 are localized within this domain (residues 564-698) (**Fig. 1A**). Moreover, EC6 is most intolerant to genetic variation in the general population, with missense tolerance ratios (MTR)^34^ in the 5^th^ percentile (**Fig. 2B**).

**Figure 2.**
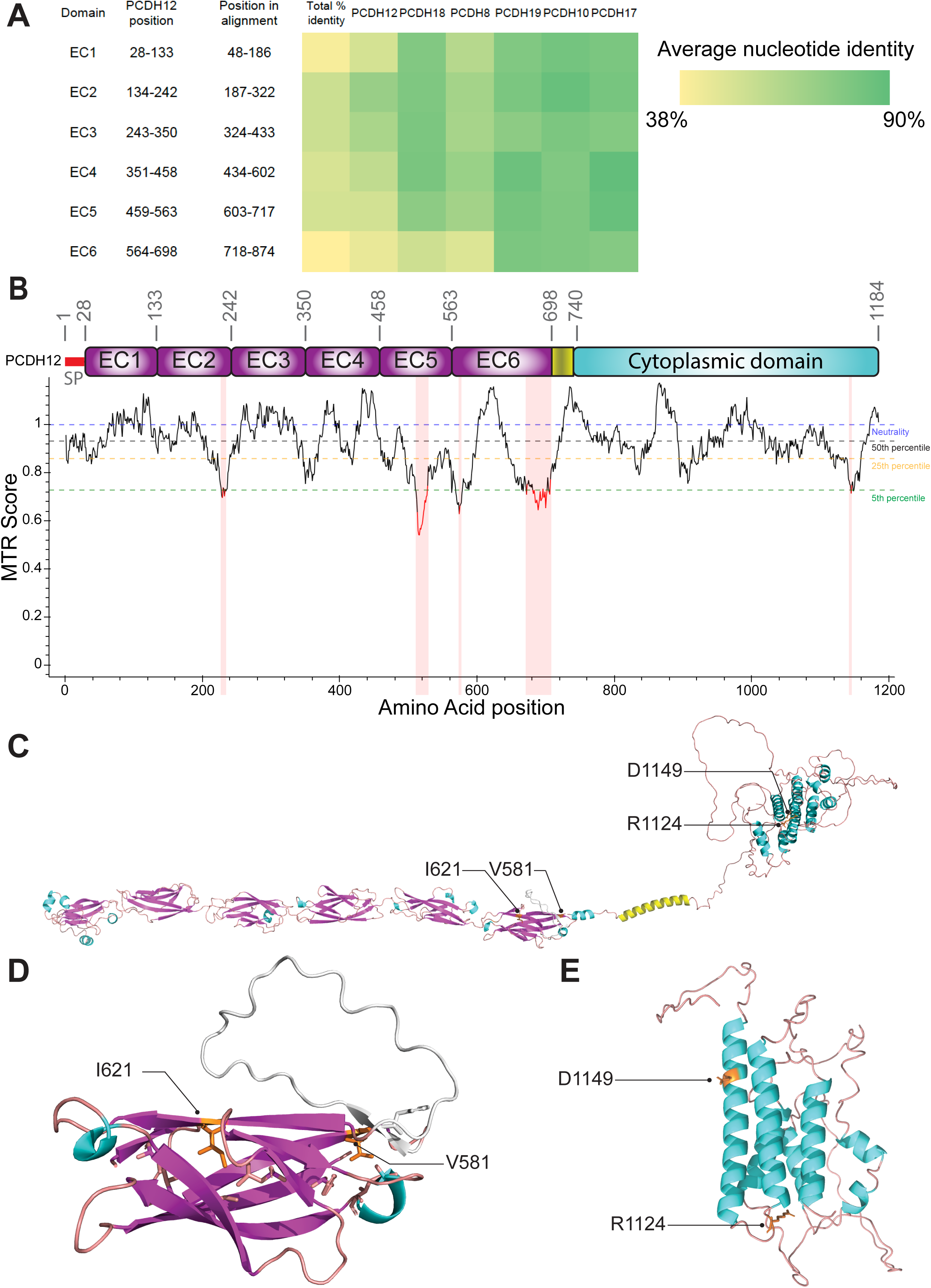
Alignment, 3D modelling, and intolerance analysis of PCDH12. (**A**) Heatmap of the mean percentage identity of each individual PCDH extracellular (EC) domain based on sequences collected from 20 phylogenetically representative vertebrates. (**B**) Graph representing the Missense Tolerance Ratio (MTR) Score along the amino acid position in PCDH12. This score measures the intolerance of a region to missense variation with 5% representing the cutoff for more intolerant segments.^34^ (**C**) AlphaFold 3D structure model of human PCDH12. (**D**) Close-up view of the EC6 domain predicted in (**C**) with location of residues Val581 and Ile621 highlighted. (**E**) Close-up view of PCDH12 predicted intracellular domain with residues Arg1124 and Asp1149 highlighted.

The missense variants p.Val581Gly and p.Ile621His surround a specific insertion in PCDH12 EC6 (residues 586-614, composed of a small β-strand and a loop) between strand A and strand B (**Fig. 2C and D, grayed region**).^35^ Using DDMut, a tool predicting the variant effects on protein stability using deep learning, both p.Val581Gly and p.Ile621His are predicted as highly destabilizing the structure (with a ΔΔG stability change value of -2.9 kcal/mol). Val581 is on the membrane-side of the Greek key beta-barrel, at the C-terminal part of the A-strand and establishes an important hydrophobic interaction network (Ile666, Phe694, Ala662 and Leu587). When replacing the valine with glycine, this hydrophobic network is lost, destabilizing the structure. Ile621, localized at the second middle of the B-strand, points towards the center of the barrel and is crucial for maintaining the hydrophobic interaction network stabilizing the barrel (with residues Val565, Val677, Leu649, and Ile675). Replacing the isoleucine with a histidine strongly destabilizes this hydrophobic network.

Residues Arg1124 and Asp1149 are localized at the C-terminal part of the intracellular domain (**Fig. 1A**), for which the structure prediction is of low accuracy. Interestingly, they are localized in a structured region predicted as an α-helix-bundle (**Fig. 2C and E**). In agreement with the low accuracy of the structure prediction, DDMut predicts both variants p.Arg1124Cys and p.Asp1149Asn as destabilizing slightly the structure (with a ΔΔG stability change value of 0.2 and 0.5 kcal/mol, respectively). P.Arg1124 is localized at the C-terminal part of an α-helix that is conserved in PCDH18.

Overall, the *in-silico* predictions strongly suggest that extracellular missense variants p.Val581Gly and p.Ile621His negatively affect PCDH12 function. The impact of *PCDH12* cytoplasmic variants p.Arg1124Cys and p.Asp1149Asn remains elusive at this stage owing to low protein prediction accuracy. However, it is likely that both intra- and extracellular variants alter PCDH12 function similarly to the reported biallelic nonsense variants. Otherwise, we would expect the mothers of families 3 and 4 to exhibit overlapping clinical phenotypes.

### Novel *PCDH12* pathogenic missense variants do not affect protein localization and expression

To gain further insight into the molecular consequences of the four missense variants p.Val581Gly, p.Ile621His, p.Arg1124Cys, and p.Asp1149Asn on PCDH12 structure and function, we transfected HEK293T cells with a C-terminal mCherry fusion of WT or variant PCDH12 proteins. As anticipated, the mCherry-WT PCDH12 fusion protein was predominantly localized in the cytoplasm near the cell membrane, while the mCherry alone exhibited a more diffused fluorescence distribution throughout the cells (**Fig. 3A, left panels**). We also noticed a somewhat polarized accumulation of WT PCDH12-mCherry signal, reminiscent of cadherin intracellular retention in organelles.^36^ PCDH12 localization appeared unaltered by any of the four missense variants (**Fig. 3A, middle and right panels**). Western blot analyses of transfected HEK293T cell lysates showed that the four variants did not affect the stability of PCDH12 proteins (**Fig. 3B and C**). We also confirmed these results in HeLa cells (**Supplementary Fig. 2**). The localization of PCDH12-WT was predominantly peri-nuclear, with some cytoplasmic presence and localization at the cell membrane. No significant differences were observed in the localization patterns between the WT and variant PCDH12 proteins (**Supplementary Fig. 2**). Altogether, these data confirm that the four new variants do not impact abundance or cellular localization of PCDH12 as part of a putative LoF mechanism.

**Figure 3.**
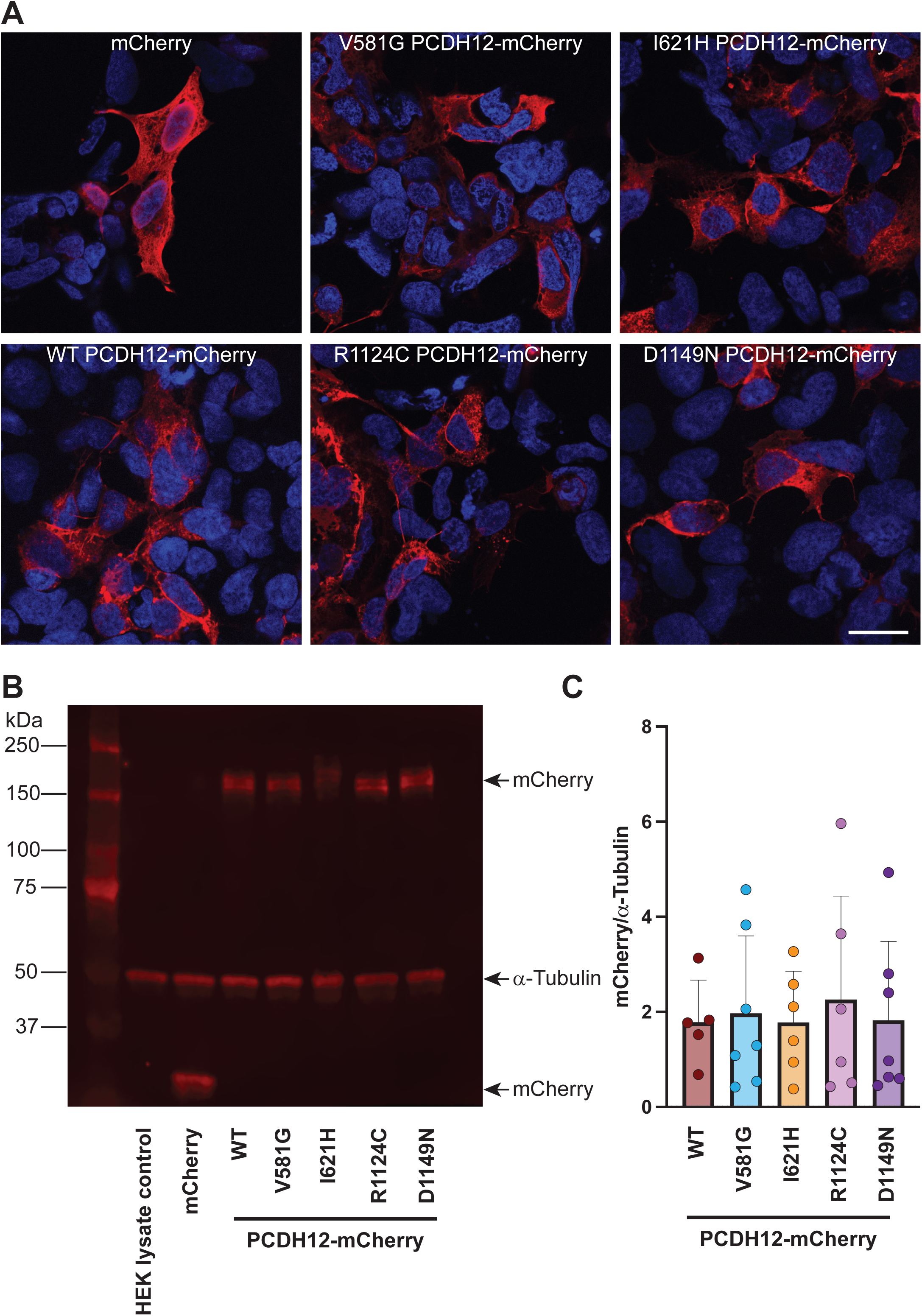
Subcellular localization and expression of PCDH12 and pathogenic missense variants. (**A**) Confocal images of HEK293T cells transfected with plasmids expressing control mCherry (top left), full-length WT PCDH12 fused to mCherry, (bottom left), p.Val581Gly PCDH12-mCherry (top middle), p.Ile621His PCDH12-mCherry (top right), p.Arg1124Cys PCDH12-mCherry (bottom middle), and p.Asp1149Asn PCDH12-mCherry (bottom right). (**B**) Representative western blot showing mCherry expression in whole cell lysates of HEK293T cells transfected with the constructs described in (**A**). A nontransfected HEK293T cell lysate was used as negative control. α-Tubulin was used as a loading control. (**C**) Quantification of mCherry signal intensity normalized to α -Tubulin in western blots of WT (*n*=5), p.Val581Gly (*n*=7), p.Ile621His (*n*=6), p.Arg1124Cys (*n*=6), and p.Asp1149Asn PCDH12-mCherry (*n*=7) expressing HEK293T cell lysates. Data are shown as mean + SD; dots represent one independent experiment. Ordinary one-way ANOVA followed by Holm-Sidák’s multiple comparisons test. Scale bar = 25 µm.

**Figure 4.**
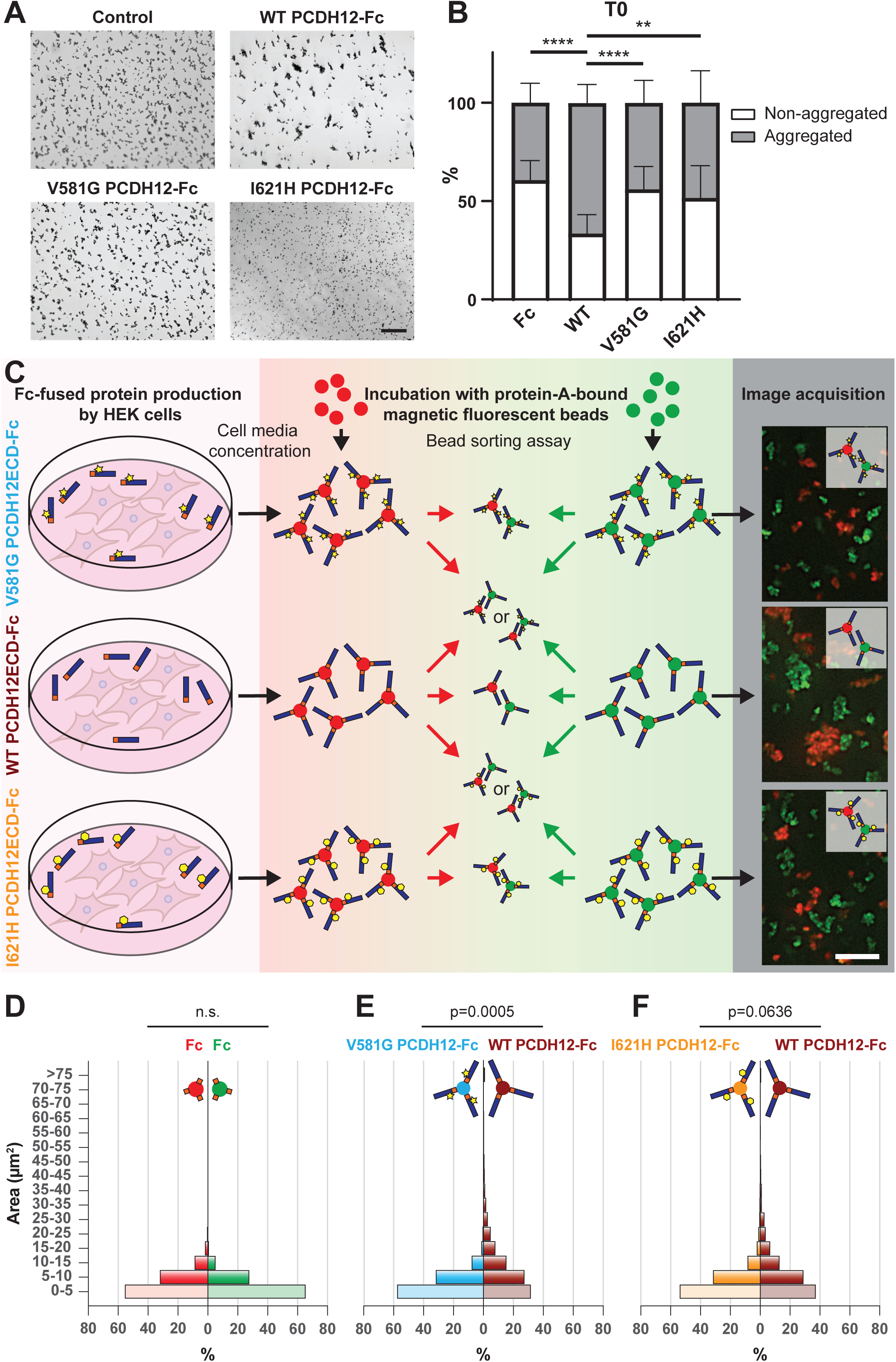
EC6 pathogenic variants negatively affect PCDH12-mediated adhesion. (**A**) Brightfield images of Protein A magnetic beads coated with the Fc domain (upper left panel), the ectodomain of WT (upper right panel), p.Val581Gly (lower left panel), and p.Ile621His (lower right panel) PCDH12 fused to Fc. Scale bar = 100 μm. (**B**) Stacked bar plot summarizing the percentage of aggregated (grey) and non-aggregated (white) beads under the four different conditions shown in (**A**) right after exposure to calcium (Time T=0). Data are shown as mean percentage + SD. *n*=14 (Fc), 31 (WT PDH12), 19 (p.Val581Gly PCDH12), and 18 (p.Ile621His). *p*<0.0001 (Fc vs. WT PCDH12 and WT vs. p.Val581Gly PCDH12); *p*=0.0016 (WT PCDH12 vs. p.Ile621His PCDH12). Brown-Forsythe and Welch ANOVA tests followed by Dunnett’s T3 multiple comparisons test. (**C**) Schematic of the fluorescent bead aggregation experimental workflow. Soluble Fc-fused WT, p.Val581Gly, and p.Ile621His PCDH12_EC_ were produced in HEK293T cells, concentrated, and bound to either green or red Protein A fluorescent magnetic beads. Green and red populations were mixed in equal amount to assess bead aggregation. Scale bar = 10 μm. (**D-F**) Distribution pyramid charts summarizing the size distribution of the bead aggregates in control (**D**), WT versus p.Val581Gly PCDH12_EC_ (**E**), and WT versus p.Ile621His PCDH2_EC_ (**F**) conditions. Fisher’s exact test.

### Extracellular *PCDH12* pathogenic variants impair adhesion

PCDHs’ extracellular domain is known to mediate homophilic trans-interactions. These interactions involve the antiparallel dimerization of ectodomains EC1 to EC4.^37^ Although variants p.Val581Gly and p.Ile621His are located on EC6 and thus not involved in the direct physical trans-contact between proteins, they are predicted to be highly destabilizing. This could lead to disruption of PCDH12 structure or trans dimers stability, and subsequently adhesion impairment. To assess whether the extracellular missense variants disrupt the adhesive potential of PCDH12 extracellular domain (PCDH12_EC_), we performed a bead aggregation assay using extracellular WT, p.Val581Gly, or p.Ile621His PCDH12 fragments fused to Fc (**Supplementary Fig. 3A**). Upon exposure to calcium, only WT-PCDH12_EC_ led to immediate aggregation, reaching 66.34 ± 9.53% of aggregates >5-µm^2^ shortly after calcium treatment (**Fig. 4A and B**), which was sustained for at least 30 minutes (56.61 ± 8.32% >5-µm^2^ aggregates) (**Supplementary Fig. 3B**). Val581Gly-PCDH12_EC_ and Ile621His-PCDH12_EC_ showed significantly lower aggregation rates similar to the Fc control (44.00 ± 11.46%, 48.35 ± 16.33%, and 39.37 ± 9.98% at T0, respectively) (**Fig. 4B**) that didn’t change even after 30 minutes (32.89 ± 11.88%, 39.22 ± 15.76%, and 34.83 ±11.04% aggregation, respectively) (**Supplementary Fig. 3B**). This result suggested that the PCDH12 extracellular missense variants negatively affect homophilic adhesion.

We next assessed whether the WT-PCDH12_EC_, Val581Gly-PCDH12_EC_, and Ile621His-PCDH12_EC_ proteins influence each other’s binding abilities. We generated two types of beads bound to each protein: one population that fluoresces green and another that fluoresces red (**Fig. 4C**). PCDH12_EC_ proteins bound to either green or red beads had similar aggregate size distributions (**Supplementary Fig. 3C-E**), with WT-PCDH12_EC_ beads generating large aggregates (**Supplementary Fig. 3C**). Beads bound to PCDH12_EC_ missense variants did not generate large aggregates (**Supplementary Fig. 3D and E)** and no difference was observed when comparing their distribution to the Fc control (**Fig. 4D)**, confirming the findings that p.Val581Gly and p.Ile621His substitutions impair PCDH12-mediated adhesion. Interestingly, when mixing WT PCDH12_EC_- with p.Val581Gly PCDH12_EC_- beads, which mirrors the heterozygosity for the extracellular domain in our patients, we observed a difference in the bin distribution between the two populations, regardless of the bead fluorescence (*p* = 0.0005) (**Fig. 4E**). On the contrary, mixing WT PCDH12_EC_- with Ile621His PCDH12_EC_-beads did not result in different distributions (*p* = 0.0636) (**Fig. 4F**). Our results suggest that co-existence of a PCDH12_EC_ missense variant copy does not affect the adhesive potential of the WT-PCDH12_EC_ copy. However, the presence of a WT copy increases the aggregation potential of Ile621His-PCDH12_EC_, which could account for the different phenotypic presentation between patients from family 3 (p.Val581Gly) and 4 (p.Ile621His).

Immunoprecipitation assays confirmed the homophilic interaction in WT PCDH12 and also showed evidence of p.Val581Gly and p.Ile621His interaction with WT PCDH12 (**Supplementary Fig. 4**). This suggests that the presence of these extracellular missense variants does not inhibit homophilic binding but rather decreases its efficiency compared to that of PCHD12 WT-WT proteins. Interestingly, even missense variants previously published and located in the EC1-EC4 domain do not prevent binding to WT PCDH12, nor missense variants located in the intracellular domain (**Supplementary Fig. 4**).

### *PCDH12* pathogenic variants do not affect the F-actin cytoskeleton

The results from the aggregation assays demonstrated adhesion but not inhibition defects. However, these findings alone do not fully explain the phenotypes observed in the affected patients. If WT/variant heterozygosity in the PCDH12 extracellular domain were sufficient to cause NDDs, we would expect parents and siblings carrying only one variant allele to be affected as well. Therefore, we hypothesized that a complementary mechanism involving both intra- and extracellular variants may contribute to pushing the phenotype toward NDDs.

A characteristic feature of δ2-PCDHs is that they carry the <u>W</u>RC Interacting <u>R</u>eceptor <u>S</u>equence, or WIRS, in their intracellular domain. This interaction potentially links PCDH12 (WIRS located at p.Phe1087-Lys1092) to the WRC and by extension to the actin cytoskeleton.^33^ Previous work showed that *PCDH12*-KO neural progenitor cells (NPCs) displayed a decreased WAVE1 recruitment at the plasma membrane compared to WT NPCs.^13^ We investigated if the *PCDH12* missense variants could affect the organization of the actin cytoskeleton. In high-confluency control cell cultures, the cells expressing the mCherry tag form dense actin filament network at cell-cell contacts (**Fig. 5A**) while at low confluency, F-actin is strongly expressed in cell protrusions (not shown). The WT-PCDH12 expression did not induce changes in actin filament organization, and level of expression did not correlate with higher or lower actin filament density (**Fig. 5B**). At low confluency, WT-PCDH12 expressing cells were still able to extend lamellipodia-like structures where the protocadherin strongly colocalized with F-actin (not shown). Similarly, no change in cell morphology and actin filament density is observed for highly confluent cultures when expressing PCDH12 variants p.Val581Gly, p.Ile621His, p.Arg1124Cys, or p.Asp1149Asn (**Fig. 5C-F**). Overall, the PCDH12 expression did not influence F-actin distribution in all our conditions. WAVE1 labeling yielded the same results; the presence of WT, p.Val581Gly, p.Ile621His, p.Arg1124Cys, or p.Asp1149Asn PCDH12 have no obvious effect on the localization and density of the cytoskeleton-associated protein (**Supplementary Fig. 5**).

**Figure 5.**
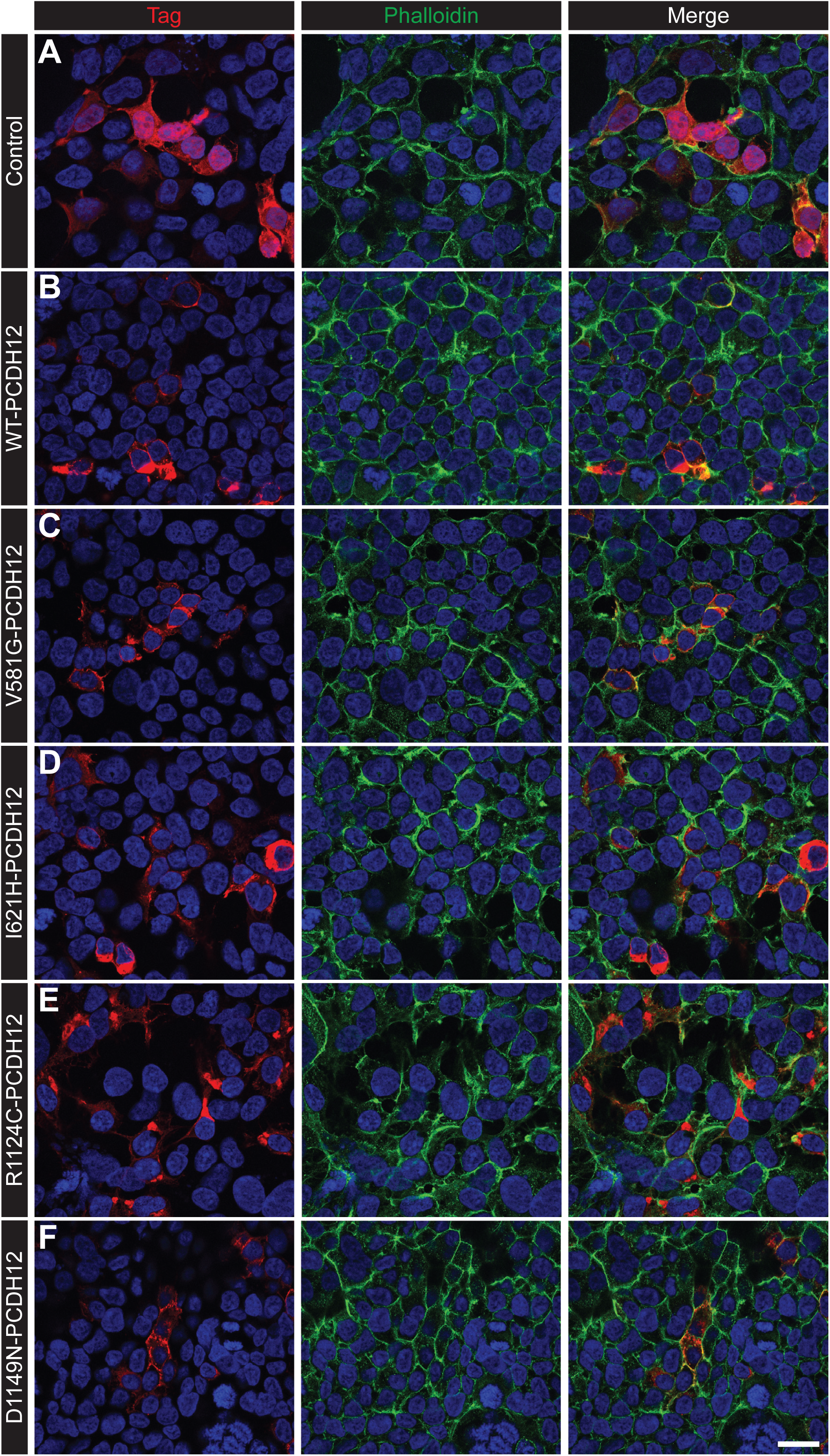
Actin cytoskeleton organization in HEK293T cells expressing PCDH12. (**A-F**) Representative confocal images of co-immunostaining of mCherry (red) alone (**A**) or C-terminally fused to WT- (**B**), p.Val581Gly- (**C**), p.Ile621His- (**D**), p.Arg1124Cys- (**E**), or p.Asp1149Asn-PCDH12 (**F**) and phalloidin (green) as a marker of filamentous actin. Nuclei were labelled with Hoechst 33342 (blue). Scale bar = 20 μm.

### Effect of *PCDH12* pathogenic variants on PCDH12 interaction with PCDH19

δ-PCDHs are known to primarily form homophilic trans dimers, while heterophilic interactions occur but are much weaker.^37^ The experiments demonstrating this relied solely on the extracellular domains of δ-protocadherins. Specifically, the extracellular EC1-EC6 domain of PCDH12 has been shown to interact with the EC1-EC4 domains of PCDH10, PCDH17, and PCDH18 to a lesser extent, whereas EC1-EC4 domains of PCDH19 exhibited exclusive homophilic binding.^37^ However, when full-length δ-PCDHs are expressed, the spectrum of heterophilic binding can widen.^31,38^ This suggests that the transmembrane or cytoplasmic domains of δ-PCDHs may contribute to these heterophilic interactions. Given that 75% of known PCDH12 patients exhibit seizures (**Supplementary Table 3**), we investigated the interaction between PCDH12 and epilepsy-related PCDH19. Our immunoprecipitation experiments confirmed PCDH19 homophilic interaction (not shown)^31^ and a direct interaction between PCDH12 and PCDH19. To identify the specific region of PCDH19 responsible for this interaction, we used full-length (FL), cytoplasmic domain (CD), extracellular domain (EC), EC-transmembrane (TM) forms of PCDH19^39^ (**Fig. 6A**). Our results showed that PCDH12 co-immunoprecipitated with PCDH19-FL, PCDH19-EC, and PCDH19-EC-TM (lanes 2, 4, 5; **Fig. 6B**) but not PCDH19-CD (lane 3), confirming that PCDH12 interacts with the extracellular domain of PCDH19. Finally, we explored whether PCDH12 missense variants could disrupt this PCDH12-PCDH19 heterophilic interaction. Notably, the p.Val581Gly and p.Ile621His variants of PCDH12 appeared to reduce PCDH19 expression (lanes 3 and 6; **Fig. 6C**), while PCDH19 expression remained unaffected by the intracellular variants, p.Arg1124Cys and p.Asp1149Asn, as well as the WT-PCDH12 (lanes 2, 4, and 5; **Fig. 6C**). As a result, immunoprecipitation results showed decreased binding of the p.Val581Gly and p.Ile621His variants to PCDH19, while the p.Arg1124Cys and p.Asp1149Asn variants showed no effect (lanes 7 to 12; **Fig. 6C**).

**Figure 6.**
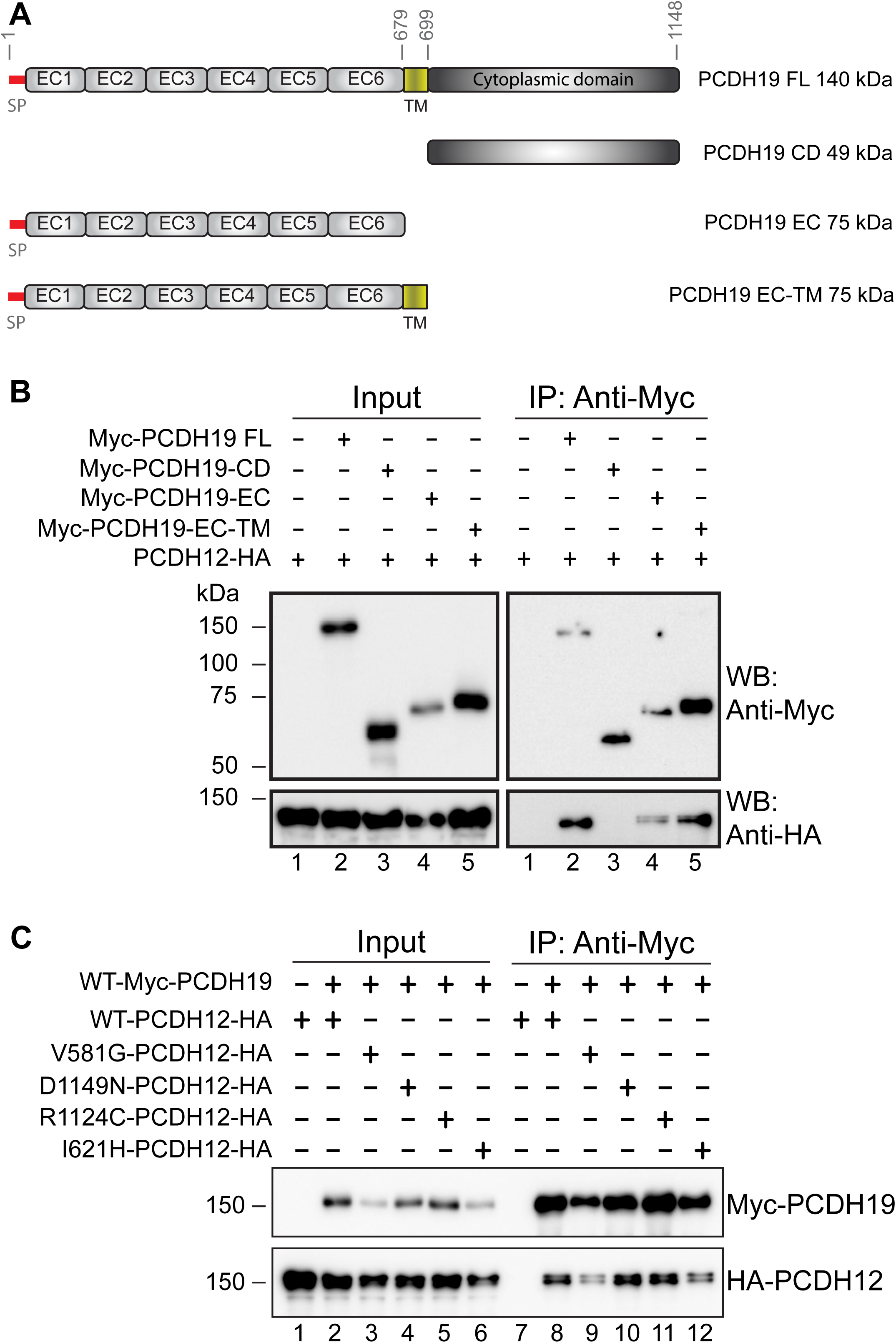
PCDH12 interacts with PCDH19. (**A**) Schematic representation of the different PCDH19 constructs used to elucidate the protein subdomain involved in the heterophilic interaction with PCDH12. SP=Signal Peptide; TM=Trans Membrane domain; FL=Full Length; CD=Cytoplasmic Domain; EC=Extra Cellular domain; EC-TM=Extra Cellular and Trans Membrane domains. (**B**) Myc-PCDH19 proteins were immunoprecipitated with anti-c-Myc magnetic beads. Inputs and immunoprecipitated samples were western blotted to detect Myc-PCDH19-FL, Myc-PCDH19-CD, Myc-PCDH19-EC, Myc-PCDH19-EC-TM, and PCDH12-HA proteins. (**C**) Combinations of Myc-PCDH19 WT and/or WT or variant HA-PCDH12 expression plasmids were transfected into HEK293T cells and the total cell lysates were immunoprecipitated using anti-c-Myc magnetic beads. Input and immunoprecipitated samples were western blotted to detect Myc-PCDH19 and HA-PCDH12 proteins.

## Discussion

To our knowledge, this study is the first to describe non-truncating missense compound heterozygous variants in *PCDH12* associated with NDD. Previous reports have identified older individuals carrying missense heterozygous variants localized in both the extracellular and intracellular domains of PCDH12. Notably, none of the patients exhibited a history of neurological or psychiatric disorders; their PCDH12-related phenotype was limited to brain calcifications.^5^ Similarly, the heterozygous parents of children carrying homozygous LoF *PCDH12* variants were found to be neurotypical.^40^ This raises intriguing questions about the neurological phenotypes presented by patients with missense compound heterozygous variants, particularly as these paired variants affect *PCDH12* at different protein domains.

Our findings indicate that the p.Val581Gly and p.Ile621His variants in the extracellular domain compromise PCDH12 binding and are predicted to be highly destabilizing, while the intracellular variants p.Arg1124Cys and p.Asp1149Asn are projected to be mildly destabilizing. Given the lack of functional evidence in our study and the absence of prior reports on intracellular missense (non-synonymous) pathogenic variants validated by functional studies across the δ2-PCDHs, including PCDH19, the impact of these cytoplasmic variants remains speculative. As such, their classification as pathogenic should be approached with caution, and they should be regarded as variants of uncertain significance.

Thermodynamic models of genetic interactions suggest that compound heterozygous variants that destabilize both protein folding and binding can significantly influence the anticipated phenotype. Notably, if ligand concentration is considerably reduced, it may lead to a more severe phenotype than what would be expected if there were no heteroallelic interactions.^41^ It is possible that destabilizing PCDH12 structure impacts its ability to serve as a ligand.^13^ The extracellular variants p.Val581Gly and p.Ile621His could impact the binding of PCDH12 produced by the WT allele in compound heterozygous individuals. Intracellular variants p.Arg1124Cys and p.Asp1149Asn could affect signal transduction triggered by PCDH12 homo- or heterophilic binding. All these factors together would act synergistically to lead to NDD. PCDH12 being susceptible to genetic interactions could also explain the phenotypic diversity observed among patients (**Supplementary Table 1**).

### *PCDH12* extracellular pathogenic variants and adhesion

The precise mechanisms by which the p.Val581Gly and p.Ile621His variants located in the PCDH12 extracellular domain disrupt adhesion remain unclear. The design of the bead aggregation assay indicates that any results obtained are directly related to PCDH12 trans-interactions. PCDH12, like the other δ2-PCDHs, engages in trans-interactions through direct contact between its EC1-EC4 domains in an anti-parallel arrangement.^37,42^ So how do variants in the EC6 domain interfere with PCDH12 trans-interactions? Our data show that these disruptions do not result from changes in PCDH12 expression levels. One plausible hypothesis is that these variants induce a conformational change that masks binding sites located within the EC1-EC4 domains, thus disrupting trans-interaction. Alternatively, there may be a reduction in the electrostatic attraction between PCDH12 proteins. Although the physical binding primarily involves the EC1-EC4 domains, even minor changes to residues on the periphery of the binding interface can significantly disrupt interactions, as electrostatic forces can influence interactions over considerable distances.^43^ For instance, the substitution of the hydrophobic isoleucine at residue 621 with the hydrophilic histidine, or the replacement of the large hydrophobic valine with a small glycine, could destabilize the structure. These changes may introduce a buried hydrophilic surface and/or decrease hydrophobic interactions with adjacent amino acids, both of which are known to diminish protein-protein binding affinities and overall interaction stability.^43,44^

### *PCDH12* pathogenic variants and the cytoskeleton

Although the structure prediction for the intracellular domain of PCDH12 shows very low accuracy (pLDDT < 50) (Fig. 2C, E), several key observations can be drawn from previous research. First, the cytoplasmic domain of PCDH12 is susceptible to proteolytic cleavage.^45^ As demonstrated in studies involving PCDH19, such cleavage can lead to the nuclear translocation of its intracellular domain, thereby influencing transcriptional activity.^46^ Second, like all δ2-PCDHs, PCDH12 contains a WIRS that facilitates the connection of its cytoplasmic tail to the actin cytoskeleton through the WRC. Specifically, PCDH10, PCDH12, and PCDH19 enhance WRC activity, whereas PCDH17 acts as an inhibitor.^33^

In our study, however, we were unable to establish a link between the functional consequences of either the intracellular variants p.Arg1124Cys and p.Asp1149Asn or the extracellular variants p.Val581Gly and p.Ile621His, and any alterations in F-actin cytoskeleton organization. This may be attributed to the limitations of our experimental model. We overexpressed PCDH12 in cells that naturally lack its expression while still possessing all WRC components. It is plausible that the expression of PCDH12 was not sufficient to significantly impact the actin cytoskeleton dynamics compared to other endogenous WRC partners in HEK293T cells. Working with neuronal models derived from patient-induced pluripotent stem cells could address this issue.

### Structural ocular malformations expand the phenotypic spectrum of *PCDH12*-associated disease

Ocular involvement in individuals with biallelic pathogenic *PCDH12* variants is predominantly characterized by retinal vascular pathology, most consistently described as exudative vitreoretinopathy or FEVR-like disease. Reported lesions include peripheral retinal avascularity, abnormal neovascularization, lipid exudation, vessel dragging, fibrosis, and tractional changes, all of which can lead to progressive visual impairment.^7,8,10^ Significant intrafamilial variability has been observed, with some individuals experiencing mild retinal changes while others suffer from profound vision loss. ^10^ These findings align with the established role of PCDH12 as an endothelial protocadherin involved in cell–cell adhesion and angiogenic remodeling, providing a biological rationale for how loss of function may disrupt retinal vascular development.^8,10^

In contrast, Family 4 presents exclusively with primary structural ocular malformations. The female proband exhibits bilateral microphthalmia and sclerocornea, while her affected brother has unilateral microphthalmia, with no signs of retinal vascular disease. Murine knockout data highlight PCDH12 importance in maintaining ocular tissue. Specifically, the *Pcdh12*-KO mouse model is associated with cataracts, and improper lens development can result in severe ocular malformations, such as microphthalmia. The presence or absence of microphthalmia or sclerocornea in mice could be attributed to species-specific compensatory mechanisms, differences in, developmental timing, or limitations in standard methods for assessing anterior segment anomalies. Importantly, genes responsible for cataracts in mice often lead to microphthalmia or other severe structural eye malformations in humans, indicating species-specific differences the expression of ocular developmental genes (e.g., *FOXE3, CHX10*, crystallins).^47^ Furthermore, the lens phenotype in the *Pcdh12*-KO mice is more frequently observed in early adult females than in males (https://www.mousephenotype.org), suggesting a sex-biased ocular effect. This is also reflected in Family 4, where the female proband exhibits a more severe phenotype than her brother, despite both carrying the same compound heterozygous *PCDH12* variants. Additional cases are needed to ascertain whether this difference is biologically significant or merely coincidental.

*PCDH12* is expressed in both neural progenitor and endothelial cells during embryogenesis, suggesting a dual role in both neural and vascular development. In the eye, PCDH12 may impact early neuroepithelial patterning, lens morphogenesis, and formation of the hyaloid and retinal vasculature. Disruptions to PCDH12 can therefore lead to both structural ocular malformations and retinal vascular defects, providing a mechanistic explanation for the spectrum of phenotypes observed in humans. Mouse studies further support the conserved functions of PCDH12 in preserving ocular tissue integrity and coordinating neurovascular development.

### Heterophilic interactions among δ2-protocadherins

The interaction between PCDH12 and PCDH19 represents a novel finding. However, the precise nature of this heterophilic interaction remains to be elucidated. Although homophilic binding is the predominant mode of adhesion, δ2-PCDHs can exhibit trans heterophilic interactions.^37^ Despite low conservation of cis interaction surfaces in δ-PCDHs compared to clustered PCDHs,^37^ there is evidence suggesting that PCDH19 can form cis heterophilic interactions with other δ-PCDHs.^31,38^

It is noteworthy that *PCDH12* extracellular variants are located within the EC6 domain, which is the cadherin domain involved in cis heterophilic adhesion in clustered PCDHs.^30,48^ In our study, we exogenously expressed PCDH12 and PCDH19 on the cell membrane, thus the observed binding between PCDH12 and the extracellular domain of PCDH19 could arise from either trans or cis interactions. Past research in animal models indicates that neurons can co-express multiple δ-PCDHs.^49,50^ This raises interesting questions about which areas of the developing human brain or cell types, co-express PCDH12, PCDH19, and other non-clustered δ-PCDHs. Understanding these co-expression patterns could provide valuable insights into the functional relevance of such interactions, enhancing our comprehension of the roles of non-clustered δ-PCDHs in cell-cell adhesion, communication, and signal transduction during brain development. *PCDH19* is one of the most relevant genes in epilepsy. Excluding the individuals of this study, 75% of *PCDH12* patients experienced seizures, with a distribution of 42% females to 58% males. This contrasts sharply with the pronounced female predominance observed in *PCDH19* patients.^51–53^ Thus, the functional significance of the PCDH12-PCDH19 interaction remains to be investigated.

Further research is needed to elucidate PCDH12’s specific role in brain and visual development, along with inclusion of more patients to examine the complete spectrum of clinical presentations and genotype-phenotype associations.

## Data Availability

All data produced in the present study are available upon reasonable request to the authors.

## Acknowledgements

We thank patients and families for their participation in this research study. We thank Dr. Talia Lerner for sharing imaging equipment (BZ-X, Keyence) and Alison Gardner for help with RNA_seq analyses. Confocal imaging work was performed at the Northwestern University Center for Advanced Microscopy (RRID: SCR_020996) generously supported by NCI CCSG P30 CA060553 awarded to the Robert H Lurie Comprehensive Cancer Center.

## Supplementary material

Supplementary material is available online.

