## Supplementary figures and images for "Novel *PCDH12* pathogenic missense variants cause neurodevelopmental disorders with ocular malformation"

### Supplementary Fig. 1

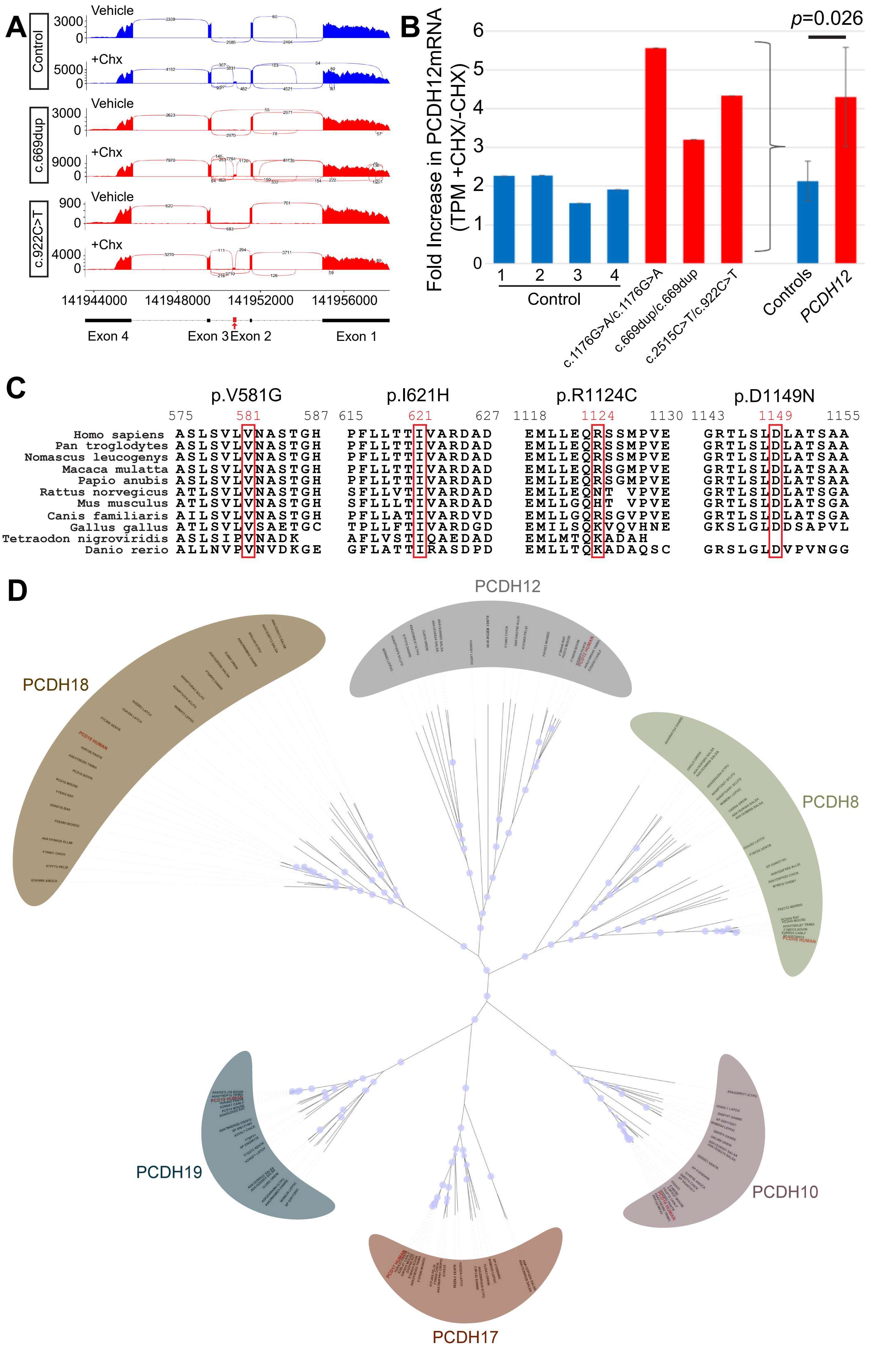


SUPPLEMENTARY FIGURE 1

### Supplementary Fig. 2

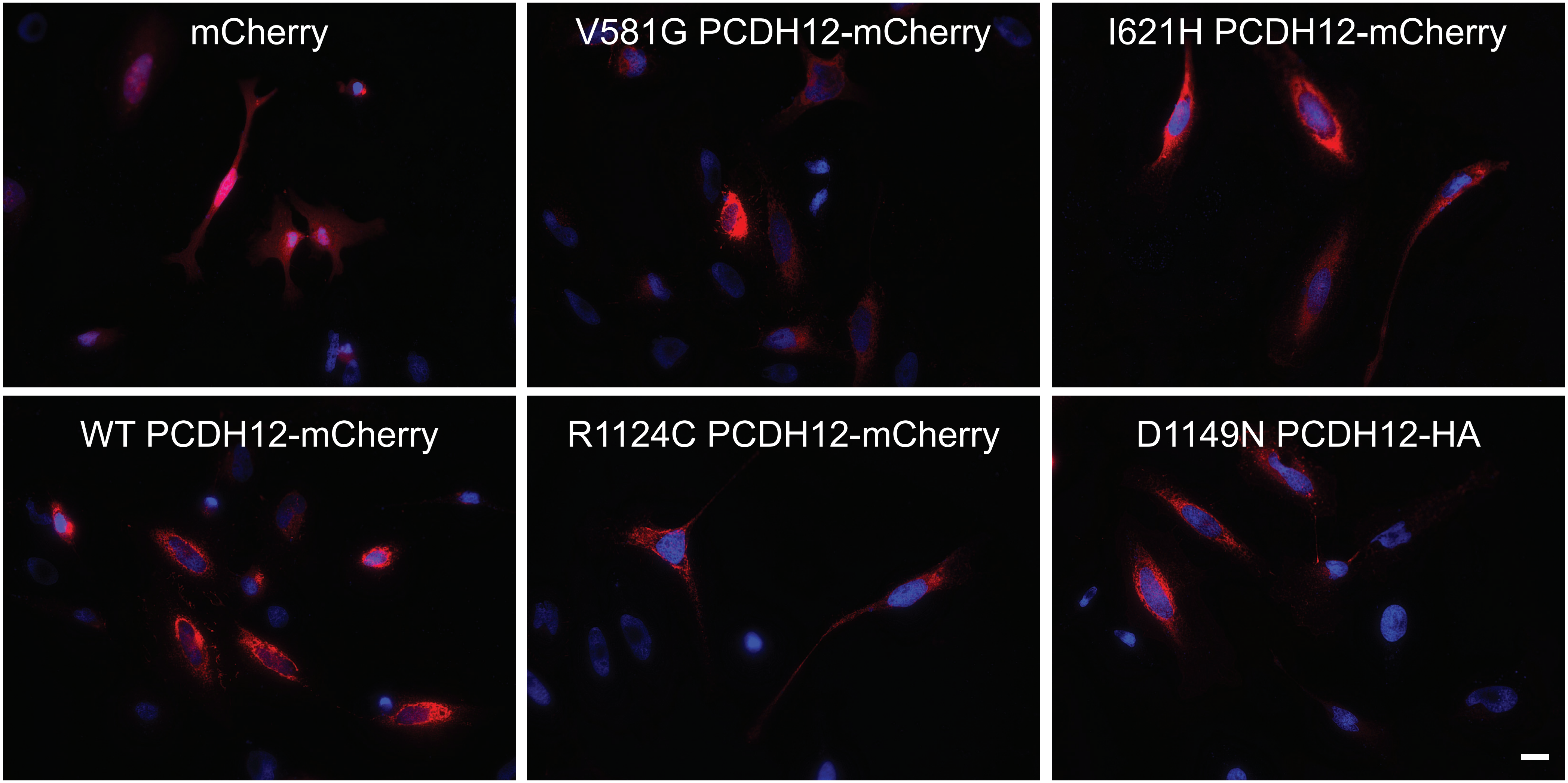


**SUPPLEMENTARY FIGURE 2**

### Supplementary Fig. 3

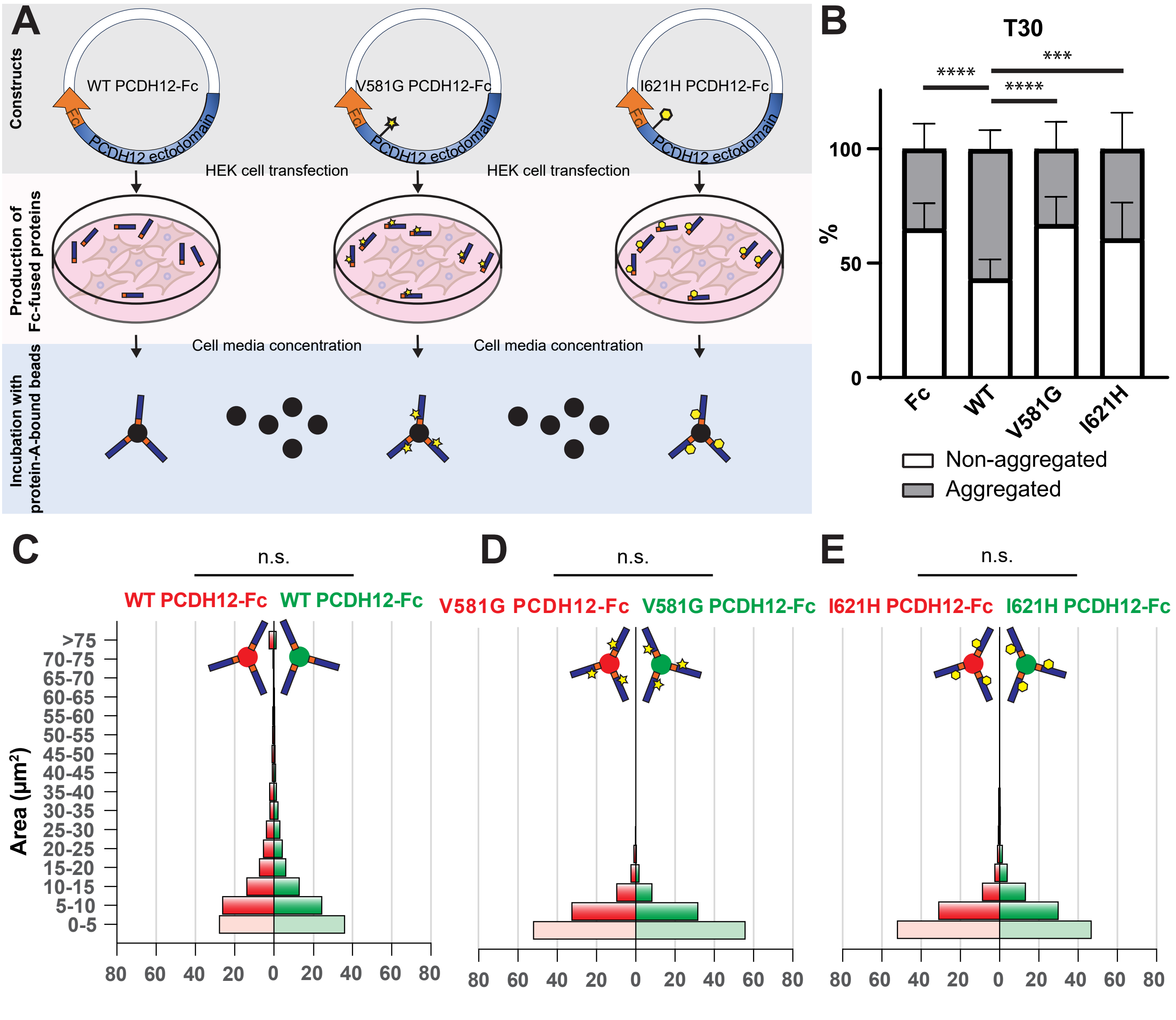


SUPPLEMENTARY FIGURE 3

### Supplementary Fig. 4

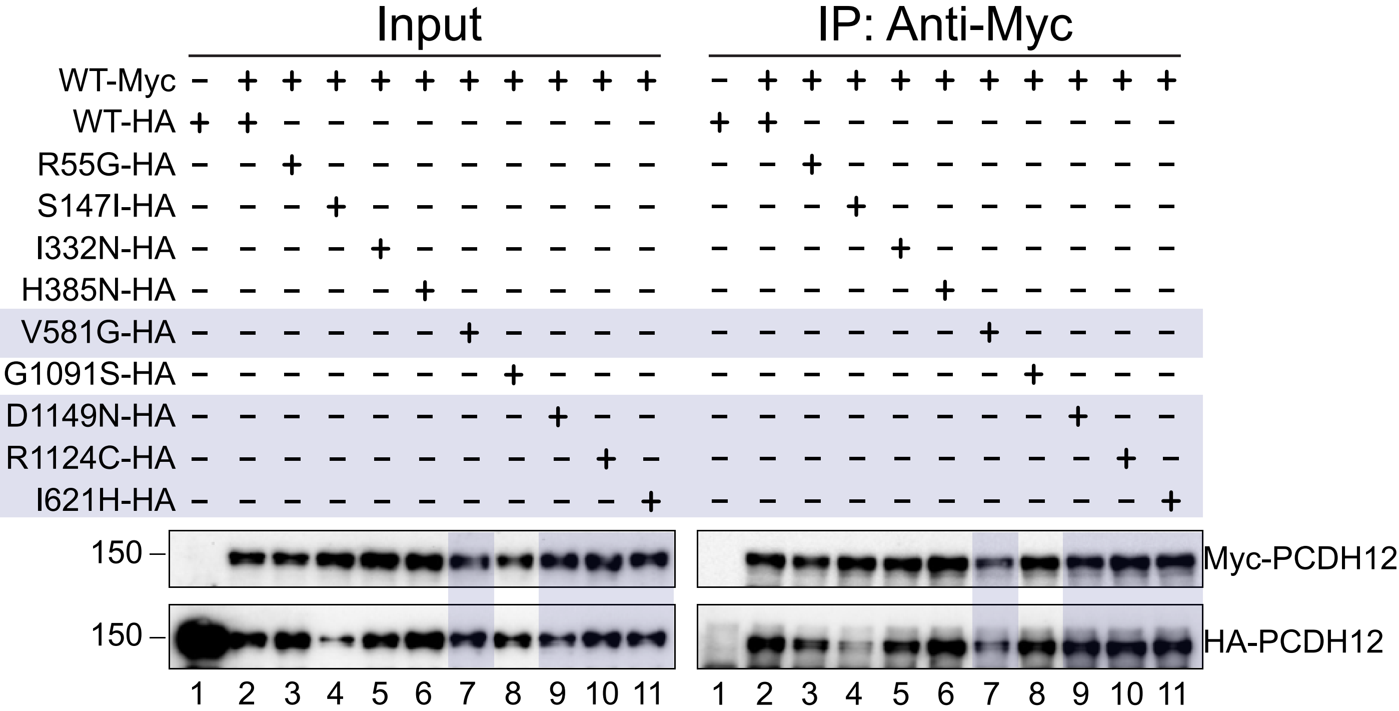


**SUPPLEMENTARY FIGURE 4**

### Supplementary Fig. 5

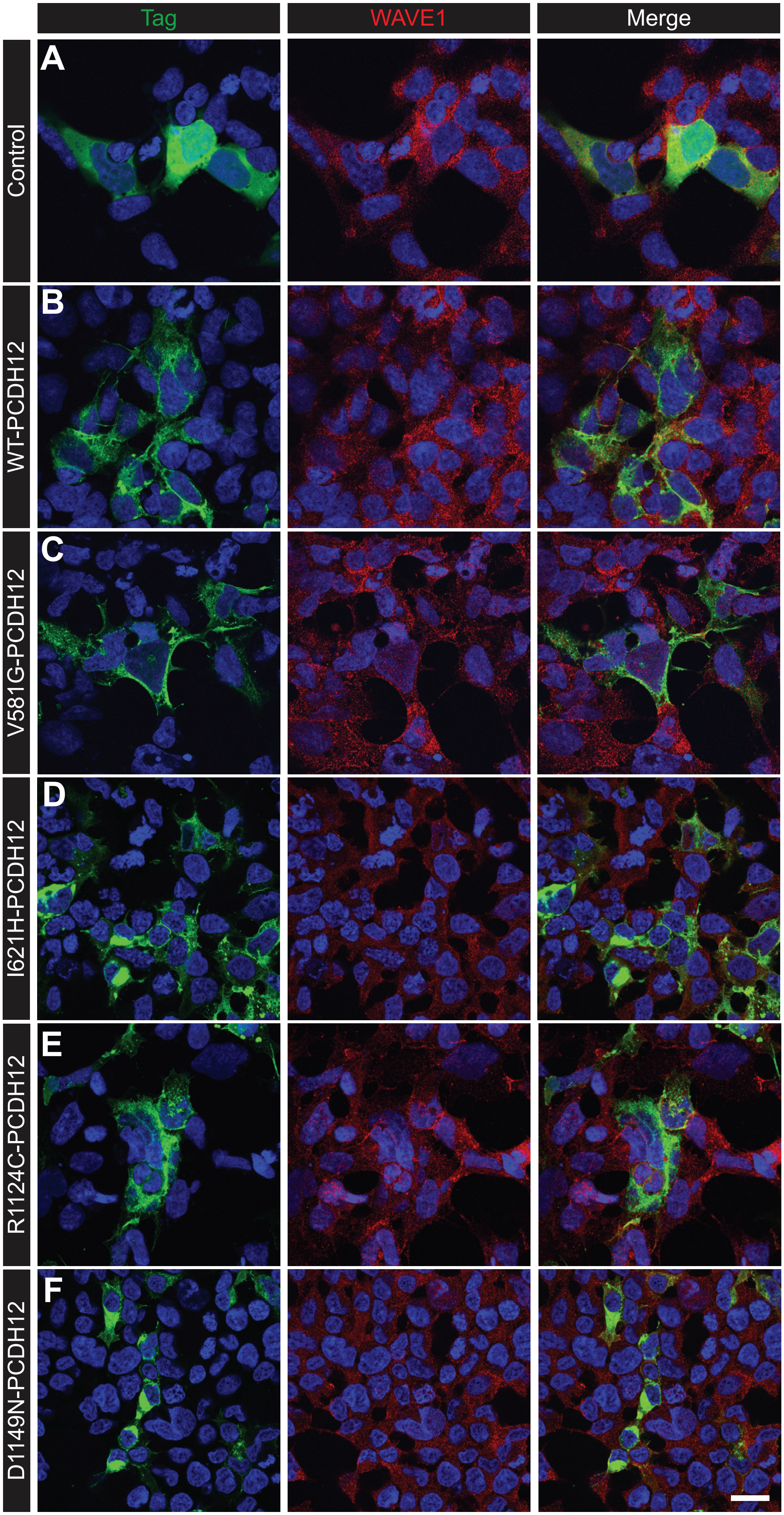


**SUPPLEMENTARY FIGURE 5**
