## Supplementary Legends for "Novel *PCDH12* pathogenic missense variants cause neurodevelopmental disorders with ocular malformation"

**Supplementary Figure Legends**

**Supplementary Figure 1. Bi-allelic pathogenic variants in *PCDH12* lead to neurodevelopmental phenotypes.** *PCDH12* truncating variant mRNAs are regulated by nonsense-mediated mRNA decay (NMD). We isolated human dermal fibroblast (HDFs) from healthy individuals (control, *n*=4; 2 males, 2 females) and those with *PCDH12* truncating variants (*n*=3; including homozygous variants NM_016580.4: c.1176G>A (p.Trp392Ter); c.669dup (p.Lys224GlnfsTer16); and compound heterozygous (Individual 2-1, Table 1; c.922C>T (p.Arg308Ter) / c.2515C>T (p.Arg839Ter)). Since PCDH12 is not expressed in HDFs, we induced *PCDH12* mRNA expression *via* gene transactivaton.^32^ Two days post- transactivation, cells were treated with or without cycloheximide (CHX) to inhibit NMD. RNA was isolated for sequencing, revealing an average PCDH12 expression of 245 +/- 75.5 TPM (transcripts per million) across all samples. (**A**) *PCDH12* variants do not alter mRNA splicing. Sashimi plots of *PCDH12* mRNA. Red arrow denotes unannotated poison exon present in all *PCDH12* mRNA samples only in the presence of CHX. (**B**). *PCDH12* variant mRNA abundance (TPM) is increased following CHX treatment compared to controls (Student’s t-test). (**C**) All four missense variants affect highly conserved residues in PCDH12 protein across different species. (**D**) Unrooted phylogenetic tree based on the full-length sequences of δ2-protocadherins PCDH8, -10, -12, -17, -18, and -19 with 1000 bootstrap iterations.

**Supplementary Figure 2. Subcellular localization and expression of PCDH12 and pathogenic missense variants in HeLa cells.** Confocal images of HeLa cells transfected with plasmids expressing mCherry (top left) and full-length PCDH12-mCherry labelled: WT PCDH12-mCherry (bottom left), Val581Gly PCDH12-mCherry (top middle), Ile621His PCDH12-mCherry (top right), Arg1124Cys PCDH12-mCherry (bottom middle), and Asp1149Asn PCDH12-HA (bottom right). Scale bar = 20 μm.

**Supplementary Figure 3. EC6 pathogenic variants negatively affect PCDH12-mediated adhesion.** (**A**) Schematic of the bead aggregation experimental workflow. (**B**) Stacked bar plot summarizing the percentage of aggregated (grey) and non-aggregated (white) beads under the four different conditions shown in (**A**) 30 minutes after exposure to calcium (Time T=30). Data are shown as mean percentage + SD. *n*=14 (Fc), 31 (WT PCDH12), 19 (Val581Gly PCDH12), and 18 (Ile621His). *p*<0.0001 (Fc vs. WT PCDH12 and WT vs. Val581Gly PCDH12); *p*=0.0010 (WT PCDH12 vs. Ile621His PCDH12). Brown-Forsythe and Welch ANOVA tests followed by Dunn’s multiple comparison tests. (**C-E**) Distribution pyramid charts summarizing the size distribution of the bead aggregates in WT versus WT (**C**), Val581Gly PCDH12_EC_ versus Val581Gly PCDH12_EC_ (**D**), and Ile621His PCDH2_EC_ versus Ile621His PCDH2_EC_ (**E**) conditions. Fisher’s exact test.

**Supplementary Figure 4. PCDH12 missense variants do not affect PCDH12 homophilic interaction.** Combinations of Myc-PCDH12 WT and/or HA-epitope-tagged WT or variant expression plasmids were transfected into HEK293T cells and the total cell lysates were immunoprecipitated using anti-c-Myc magnetic beads. Input and immunoprecipitated samples were western blotted to detect Myc-and HA-PCDH12 proteins. Missense variants described in this study are highlighted. Data shown from one of the two independent experiments (*n*=2).

**Supplementary Figure 5. WAVE1 localization in HEK293T cells expressing PCDH12 and pathogenic missense variants.** (**A-F**) Representative confocal images of co-immunostaining of HEK293T cells transfected with plasmids expressing control GFP (green) alone (A) or C-terminally fused to full-length WT- (**B**), Val581Gly- (**C**), Ile621His- (**D**), Arg1124Cys- (**E**), or Asp1149Asn-PCDH12 and WAVE1 (red). Nuclei were labelled with Hoechst 33342 (blue). Scale bar = 20 μm.
