## Supplementary Table 1 for "Novel *PCDH12* pathogenic missense variants cause neurodevelopmental disorders with ocular malformation"

**Supplementary Table 1 Overview of published bi-allelic *PCDH12* variants and main clinical findings**

| **Variant^(Reference)^** | **Sex** | **DMJD** | **Microcephaly** | **Seizures** | **GDD** | **ID** | **ASD** | **Calcifications** | **Visual Impairment** | **Hypotonia** | **Dystonia** | **Spasticity** | **Family/patient** |
| --- | --- | --- | --- | --- | --- | --- | --- | --- | --- | --- | --- | --- | --- |
| p.R839*^(4)^ | F | Yes | Yes | Yes | Yes | N/R | N/R | N/R | Yes | Yes | Yes | No | 1-1 |
| p.R839*^(4)^ | M | N/A | Yes | N/A | N/A | N/A | N/A | N/D | N/A | N/A | N/A | N/A | 1-2 |
| p.R839*^(4)^ | M | Yes | Yes | Yes | Yes | N/A | N/A | No | Yes | Yes | Yes | No | 2-1 |
| p.R839*^(4)^ | M | N/A | N/A | N/A | N/A | N/A | N/A | N/D | N/A | N/A | N/A | N/A | 2-2 |
| p.R839*^(4)^ | F | No | Yes | Yes | Yes | Yes | N/R | No | Yes | N/R | N/R | Yes | 3-1 |
| p.R839*^(4)^ | F | No | Yes | Yes | Yes | Yes | N/R | No | Yes | N/R | N/R | Yes | 3-2 |
| p.R839*^(4)^ | M | No | Yes | Yes | Yes | Yes | N/R | No | Yes | N/R | N/R | Yes | 3-3 |
| p.R839*^(4)^ | F | No | Yes | Yes | Yes | Yes | N/R | No | Yes | N/R | N/R | Yes | 3-4 |
| p.R839*^(4)^ | F | No | Yes | Yes | Yes | Yes | N/R | No | Yes | Yes | Yes | Yes | 3-5 |
| p.R839*^(4)^ | F | Yes | Yes | N/A | N/A | N/A | N/A | N/D | N/A | N/A | N/A | N/A | 4-1 |
| p.L150Afs*11/p.S175Pfs*22^(6)^ | M | No | No | Yes | Yes | Yes | N/R | Yes | N/R | Yes | No | Yes | 5-1 |
| p.S838fs*^(3)^ | M | Yes | Yes | Yes | Yes | Yes | Yes | Yes | Yes | No | No | Yes | 6-1 |
| p.S838fs*^(3)^ | M | Yes | Yes | Yes | Yes | Yes | No | Yes | Yes | No | No | Yes | 6-2 |
| p.P922fs*^(3)^ | F | Yes | Yes | Yes | Yes | Yes | Yes | N/A | Yes | No | No | Yes | 7-1 |
| p.P922fs*^(3)^ | M | Yes | Yes | Yes | Yes | Yes | Yes | N/A | Yes | No | No | Yes | 7-2 |
| p.P922fs*^(3)^ | M | Yes | Yes | Yes | Yes | Yes | Yes | N/A | Yes | No | No | No | 8-1 |
| p.P922fs*^(3)^ | M | Yes | Yes | Yes | Yes | Yes | No | Yes | N/R | Yes | No | No | 8-2 |
| p.P922fs*^(3)^ | F | Yes | Yes | Yes | Yes | Yes | No | No | No | Yes | No | No | 9-1 |
| p.P922fs*^(3)^ | M | Yes | Yes | No | Yes | Yes | No | Yes | No | Yes | No | No | 9-2 |
| p.E280*^(3)^ | F | Yes | Yes | No | Yes | Yes | No | N/A | Yes | Yes | No | No | 10-1 |
| p.R151*^(3)^ | M | Yes | Yes | Yes | Yes | Yes | No | Yes | Yes | No | No | No | 11-1 |
| p.R151*^(3)^ | F | Yes | Yes | Yes | Yes | Yes | No | Yes | Yes | No | No | No | 11-2 |
| p.R151*^(3)^ | M | Yes | Yes | N/A | Yes | Yes | No | N/A | No | No | No | No | 11-3 |
| p.P922fs*^(3)^ | M | Yes | Yes | Yes | Yes | Yes | No | N/A | No | No | No | No | 12-1 |
| p.V354fs*^(3)^ | M | Yes | Yes | Yes | Yes | Yes | No | N/A | No | No | N/R | N/R | 13-1 |
| p.E670*^(7)^ | F | No | No | Yes | No | No | N/R | No | Yes | No | Yes | No | 14-1 |
| p.E670*^(7)^ | M | No | No | No | No | No | N/R | No | Yes | No | Yes | No | 14-2 |
| p.K224Qfs*16^(8)^ | M | N/D | Yes | N/R | Yes | N/R | N/R | N/D | Yes | N/A | No | Yes | 15-1 |
| p.V724Yfs*8^(10)^ | F | Yes | Yes | No | Yes | Yes | N/D | No | Yes | Yes | Yes | Yes | 16-1 |
| p.V724Yfs*8^(10)^ | F | Yes | Yes | No | Yes | Yes | N/D | No | Yes | Yes | Yes | No | 16-2 |
| p.W392*^(9)^ | M | No | Yes | Yes | Yes | N/R | N/R | Yes | Yes | Yes | Yes | No | 17-1 |
| p.W808*^(9)^ | F | No | Yes | Yes | Yes | N/R | N/R | No | Yes | N/R | N/R | Yes | 18-1 |
| p.W808*^(9)^ | M | No | Yes | Yes | Yes | N/R | N/R | Yes | Yes | Yes | Yes | Yes | 18-2 |
| p.Q813*^(9)^ | M | No | Yes | N/R | NR | Yes | N/R | No | Yes | No | Yes | Yes | 19-1 |
| p.Q813*^(9)^ | F | No | Yes | N/R | NR | Yes | N/R | No | Yes | No | Yes | Yes | 19-2 |
| p.R151*^(11)^ | M | Yes | Yes | No | Yes | Yes | N/R | No | N/R | Yes | No | No | 20-1 |
| p.R151*^(11)^ | F | No | Yes | No | Yes | Yes | N/R | No | N/R | No | No | No | 20-2 |
| p.R99*^(12)^ | M | Yes | Yes | Yes | Yes | Yes | Yes | No | Yes | No | No | Yes | 21-1 |
| p.R99*^(12)^ | M | No | Yes | No | Yes | Yes | No | No | Yes | No | No | Yes | 21-2 |

Abbreviations: N/A: Not Available; N/R: Not Reported; N/D: Not done; F: Female; M: Male

DMJD: Diencephalic–mesencephalic junction dysplasia; GDD: Global Developmental Delay; ID: Intellectual Disability; ASD: Autism Spectrum Disorder
