## Supplementary Table 2 for "Novel *PCDH12* pathogenic missense variants cause neurodevelopmental disorders with ocular malformation"

| **Supplementary Table 2 *In Silico* predictions for newly identified *PCDH12* variants** | | | | | | | | | | |
| --- | --- | --- | --- | --- | --- | --- | --- | --- | --- | --- |
| **HGVSc** | **HGVSp** | **gnomAD total AF (V.4.1.0)** | **SIFT** | **PPH2** | **CADD** | **ClinPred** | **M-CAP** | **Mutation Taster** | **PANTHER** | **GERP** |
| NM_016580.3(PCDH12):c.922C>T | p.Arg308* | 0.00006196 | N/A |  |  |  |  |  |  | 3.44 |
| NM_016580.3(PCDH12):c.1742T>G | p.Val581Gly | 0 | Deleterious (0.03) | Probably damaging (0.997) | 24.3 | 0.872 | 0.043 | Disease causing (p-value: 1) | Possibly damaging 0.5 | 5.38 |
| NM_016580.3(PCDH12):c.1861_1862delinsCA | p.Ile621His | 0 | Deleterious (0) | Probably damaging (0.997) | N/A | N/A | N/A |  |  |  |
| NM_016580.3(PCDH12):c.3370C>T | p.Arg1124Cys | 0.00002354 | Deleterious (0) | Possibly damaging (0.549) | 25.8 | 0.550 | 0.126 | Disease causing (p-value: 0. 834) | Probably benign 0.27 | 3.33 |
| NM_016580.3(PCDH12):c.3445G>A | p.Asp1149Asn | 0 | Deleterious (0.02) | Possibly damaging (0.707) | 25.2 | 0.930 | 0.024 | Disease causing (p-value: 1) | Possibly damaging 0.5 | 6.07 |

Abbreviations: N/A: Not Available; HGVS: Human Genome Variation Society; gnomAD: The Genome Aggregation Database; SIFT: Sorting Intolerant From Tolerant; PPH2: Polymorphism Phenotyping v2; CADD: Combined Annotation Dependent Depletion; M-CAP: Mendelian Clinically Applicable Pathogenicity; PANTHER: Protein Analysis Through Evolutionary Relationship; GERP: Genomic Evolutionary Rate Profiling
