## Supplementary Table 3 for "Novel *PCDH12* pathogenic missense variants cause neurodevelopmental disorders with ocular malformation"

**Supplementary Table 3 Summary of main phenotypes seen in individuals with biallelic *PCDH12* variants**

|  | **39 previously described cases 16F (41%), 23M (59%)** | |
| --- | --- | --- |
| **Phenotype** | **%** | **%** |
| Midbrain Dysplasia | 58 | 21/36 |
| Brain Calcifications | 33 | 9/27 |
| Microcephaly | 92 | 35/38 |
| Seizure | 75 | 24/32 |
| Global Developmental Delay | 94 | 32/34 |
| Intellectual disability | 93 | 28/30 |
| Autism Spectrum Disorder | 31 | 5/16 |
| Visual Impairment | 84 | 27/32 |
| Hypotonia | 46 | 13/28 |
